# Task-specific fine-tuning versus frontier large language models for RoB 2 risk of bias assessment: development and external benchmark evaluation of RoB-BERT

**DOI:** 10.64898/2026.11.19.26368519

**Authors:** Tyler Pitre, Jasmine Mah, Ali Azlan, David Nguyen, Dena Zeraatkar, Arnav Agarwal, Gordon Guyatt

**Author notes:** Corresponding author: Tyler Pitre, MD, PhD, Division of Respirology, Department of Medicine, University Health Network and University of Toronto, Toronto, ON, Canada.

## Abstract

**Background:** Researchers increasingly use large language models (LLMs) to assess risk of bias in randomized trials, but how task-specific fine-tuned models compare with frontier models remains uncertain.

**Objective:** To develop RoB-BERT, a fine-tuned RoB 2 assessment system, and to compare its agreement with human assessments against that of three frontier LLMs.

**Methods:** RoB 2 rates five bias domains and the trial overall as low risk, some concerns or high risk, based on answers to 22 signalling questions. RoB-BERT answers each question, quotes the supporting sentences from the trial report, and applies the official RoB 2 algorithm. We trained RoB-BERT on 5,304 reviewer-checked assessments, none from a test trial, and compared it with Claude Fable 5.1, GPT-6 Astra and Gemini 4 Argon, each given identical prompts and text, against 419 published human assessments (ROBoto2, LogiMed-RoB and Cochrane reviews) made without AI assistance. Each reference was the single published judgement for that trial, made by one assessor or agreed by two, rather than an expert reference standard, so we also report agreement between two human assessors of the same trial.

**Results:** In ROBoto2, pairs of human assessors who rated the same trial agreed on 61.2% to 64.7% of domain judgements (κ 0.15 to 0.35) and 46.2% to 50.0% of overall judgements (κ 0.15 to 0.17); across the 185 ROBoto2 trials, RoB-BERT agreed with the human judgement on 67.4% (κ 0.35) and 53.5% (κ 0.20). Across all 419 assessments, domain agreement was 67.9% (κ 0.29) for RoB-BERT, 65.0% (κ 0.24) for Claude Fable 5.1, 52.7% (κ 0.21) for GPT-6 Astra and 66.7% (κ 0.26) for Gemini 4 Argon; overall agreement was 46.1% (κ 0.16), 47.3% (κ 0.17), 31.0% (κ 0.03) and 46.5% (κ 0.14), respectively. For domains the reference rated high risk, sensitivity was 22.2% for RoB-BERT, 23.1% for Claude Fable 5.1, 41.9% for GPT-6 Astra and 24.8% for Gemini 4 Argon, with specificity of 92.5%, 92.8%, 80.0% and 95.0%. On the 52 ROBoto2 trials rated by two assessors, the second assessor’s sensitivity for the reference assessor’s high-risk domains was 23.1% (specificity 90.3%) and RoB-BERT’s was 30.8% (specificity 92.7%). RoB-BERT located 95.7% of its quotations verbatim in the report and cost US$0.005 per trial, 2% to 6% of the frontier models’ cost.

**Conclusions:** RoB-BERT agreed with single human assessors at least as closely as two human assessors agreed with each other, and matched or exceeded frontier LLMs at a fraction of their cost. The results show the poor reliability of human assessors using RoB 2 and the need for consensus ratings, with RoB-BERT potentially serving as one rater.

## Introduction

The Cochrane risk-of-bias tool for randomized trials (RoB 2), a popular option for rating risk of bias in randomized trials, requires experienced reviewers to answer 22 signalling questions across five bias domains for every outcome of interest (1, 2). In one systematic review, assessment took a mean of almost six hours of staff time per study (3), and reliability is limited even between trained assessors: in one study, agreement between assessors on the overall judgement was κ −0.15 before they used detailed implementation guidance and κ 0.42 once they had (4). As trial volume grows and living reviews shorten update cycles, the assessment step represents a bottleneck in evidence synthesis.

Large language models (LLMs) offer one strategy to address that bottleneck, and several groups have evaluated general-purpose chatbots against human RoB 2 assessments (5–8). General-purpose LLMs read long reports and write readable rationales, but their answers vary between runs, sometimes describe procedures the report does not contain, change with prompt wording, and add a non-trivial cost per trial (5, 6, 8). Smaller classification models of the BERT family (9–11) are fast and, because they choose among fixed answers rather than write text, do not invent content, but they cannot cite the sentence behind an answer and have shown limited agreement with experts (12–15).

Whether supervised fine-tuning on reviewer-audited assessments can match or surpass frontier models on a shared benchmark remains untested. A small, local, deterministic system performing comparably to hosted frontier models could reduce the cost and improve the privacy and reproducibility of large-scale evidence synthesis.

We developed RoB-BERT, a hybrid of a fine-tuned generative reader and a biomedical encoder feeding the published RoB 2 decision algorithm, and compared it with Claude Fable 5.1, GPT-6 Astra and Gemini 4 Argon on a fixed external benchmark of 570 primary human assessments under a pre-registered protocol.

## Methods

### Study design and reporting

We report this developer-led model development and external evaluation study according to TRIPOD-LLM (16). The evaluation followed the RoB-BERT Fine-Tuned Models Protocol, version 1.3, which we registered on the Open Science Framework before running any comparator (https://doi.org/10.17605/OSF.IO/47ZYX). We did not amend the protocol. We fixed reference assessments, prompts, settings, and model weights before external runs began and verified them by checksum (Supplementary Table S1).

### Development corpus

The development corpus consisted of 5,304 RoB 2 worksheets, each an outcome-level assessment of an individually randomized parallel-group trial, from four sources (Table 1): a reviewer-audited trial collection, audited assessments from 18 systematic review projects, a reviewed selection from a larger pool of draft assessments, and 16 assessments completed independently by one investigator and reserved for evaluation. We grouped worksheets by trial so that reports or outcome rows of the same trial could not fall on both sides of a split, giving 4,343 trial groups partitioned into training (4,237 worksheets), validation (519), internal test (532) and the independent investigator set (16). Before training we screened the corpus against every benchmark trial by identifier, DOI and title; two training rows matched and were removed, so no benchmark trial contributed to training or validation.

**Table 1.** Sources of the development corpus.

| Source | Worksheets supplied | Worksheets retained |
|---|---|---|
| Reviewer-audited trial collection | 3,988 | 3,986 |
| Audited systematic review assessments from 18 projects | 960 | 941 |
| Reviewed assessments selected from a 1,794-record pool | 361 | 361 |
| Independent investigator assessments reserved for evaluation | 16 | 16 |
| Total | 5,325 | 5,304 |

Worksheets are outcome-level RoB 2 assessments of individually randomized parallel-group trials. Nineteen worksheets from the systematic review projects were dropped at audit for incomplete or inconsistent answers, and two worksheets from the reviewer-audited collection were removed because their trials appear in the external benchmark.

### System architecture

RoB-BERT v6.1 has three parts (Figure 1). The reader is Qwen3.8-27B with a QLoRA adapter (17), trained in two supervised stages on the training worksheets using FlashAttention and gradient checkpointing to fit long inputs on a single accelerator (18, 19). Given the trial text, outcome and effect of interest, it emits a structured record for each of the 22 signalling questions: the answer on the five-category RoB 2 scale (yes, probably yes, probably no, no, no information), one or more verbatim quotations, and a short rationale. Decoding follows a grammar that enforces the RoB 2 question flow, including branches that depend on earlier answers, at temperature 0 with extended thinking off. Quotations are checked against the source text after generation; unlocatable quotations stay in the output but are counted as unlocated. The system normalizes the input, splits it into sections and truncates it by section priority to a 16,384-token context when the article is longer, allowing up to 3,072 new tokens for the answer.

**Figure 1.**
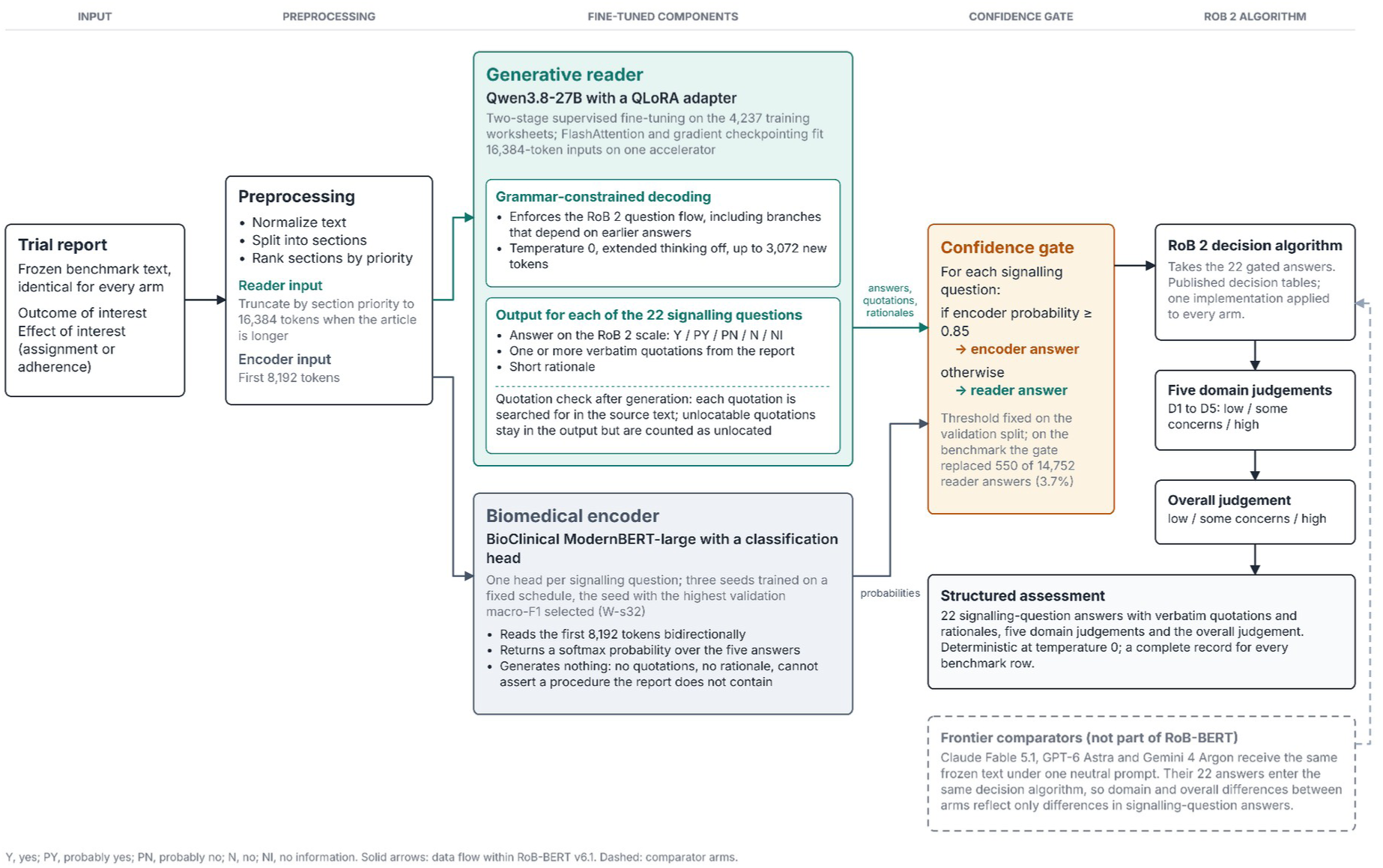
Architecture of RoB-BERT v6.1. Trial text is normalized and split into prioritized context windows for the generative reader (Qwen3.8-27B fine-tuned via QLoRA) and biomedical encoder (BioClinical ModernBERT-large). The reader generates structured answers, verbatim quotations, and rationales for 22 signalling questions. The encoder provides confidence probabilities; answers with probability ≥0.85 override the reader. Gated answers enter the RoB 2 decision algorithm to compute domain and overall judgements, alongside frontier comparators receiving identical input.

The encoder is BioClinical ModernBERT-large, a long-context biomedical encoder (14), with a classification head that predicts the answer category for each signalling question from the first 8,192 tokens of the same input. We trained three seeds on a fixed schedule and selected the seed with the highest validation macro-F1 (W-s32) before external evaluation; its outputs are unscaled softmax probabilities (15). In the combined system the encoder acts as a gate: wherever its probability for an answer reaches 0.85, that answer replaces the reader’s. We set the threshold on the validation split and did not change it afterwards.

Signalling-question answers from every AI system, including the frontier models, pass through the same implementation of the published RoB 2 decision tables to produce domain judgements and the overall judgement (2). Domain-level and overall differences between systems therefore reflect only differences in signalling-question answers, not differences in how the algorithm was applied.

### External benchmark and reference assessments

Our external evaluation used the RoB 2 AI Benchmark, version 1.0.0, a fixed, publicly available collection of RoB 2 assessments with corresponding trial texts, unchanged since its release (https://github.com/CoreSR/rob2-ai-benchmark). We assembled it from three published sources: the ROBoto2 dataset, which contains both unassisted assessments and assessments made with an LLM’s suggestions on screen (assisted) (5); LogiMed-RoB Track A (13); and Cochrane review assessments used in an earlier evaluation of GPT-4, restricted to the 41 trials with open-access full text (6). Unless otherwise specified, human assessments in this report are the unassisted assessments. The reference for each row is the single judgement recorded by the source project, made by one assessor or agreed by two; no reference was adjudicated by an independent expert panel. We verified labels and texts against published checksums before any run.

The primary population comprised the 570 primary assessments with available trial text: 185 ROBoto2 unassisted, 192 LogiMed, and 42 Cochrane rows (419 pooled unassisted rows, 418 trials), plus the 151 ROBoto2 assisted rows, which the protocol specified for separate analysis because an LLM assisted in producing their reference (Table 2). ROBoto2 rows assess the primary result at the study level; LogiMed and Cochrane rows name a specific outcome. A sensitivity population included every scored outcome row with text (1,008 rows in all: LogiMed 600, Cochrane 72, ROBoto2 336).

**Table 2.** Composition of the external benchmark (RoB 2 AI Benchmark v1.0.0).

| **Cohort** | **Source** | **Reference assessment** | **Unit assessed** | **Primary rows (trials)** | **All outcome rows** | **Signalling-question reference** |
|---|---|---|---|---|---|---|
| ROBoto2 unassisted | Hevia et al. 2025 | Human assessor without LLM assistance | Study level, primary result | 185 (185) | 185 | Yes |
| LogiMed-RoB Track A | Huang et al. 2026 | Human expert assessment | Named outcome | 192 (192) | 600 | No |
| Cochrane | Pitre et al. 2026 (open-access trials only) | Cochrane review authors | Named outcome | 42 (41) | 72 | No |
| Pooled unassisted cohorts |  |  |  | 419 (418) | 857 | ROBoto2 only |
| ROBoto2 assisted | Hevia et al. 2025 | Human assessor with LLM suggestions on screen; analyzed separately | Study level, primary result | 151 (151) | 151 | Yes |
| Total |  |  |  | 570 (569) | 1,008 |  |

Every system received the same frozen benchmark text for each row; no PDFs, registrations or protocols were supplied. Seven LogiMed trials that also appear in the Cochrane set are scored in the Cochrane set only. The protocol stated 1,001 all-outcome rows; the benchmark scorer's definition gives 1,008 and was used.

### Frontier comparators

We accessed Claude Fable 5.1, GPT-6 Astra and Gemini 4 Argon through the OpenRouter API (20) with identical settings: high reasoning effort, a 16,000-token output limit and provider defaults otherwise. Each request carried a fixed system prompt, identical for every model (checksummed and printed in the protocol) and a user message containing the RoB 2 tool block with the 22 signalling questions, the outcome, the effect of interest and the frozen benchmark text, so every system saw exactly the text given to RoB-BERT. Rows with byte-identical prompts shared one request (961 unique requests for 1,008 rows); all primary rows were unique. An output with fewer than 22 parsed answers was requested once more; a second failure remained invalid. We did not edit any output, and we recorded the time, settings, returned model and provider, token counts, billed cost, finish reason and every attempt. Preliminary runs of Claude Fable 5.1 and GPT-6 Astra on the 336 ROBoto2 rows on 12 October 2026, made with different output limits and reasoning settings while the pipeline was being built, are excluded from all comparisons; we report their agreement with the protocol runs only as a measure of run-to-run variation (Supplementary Table S2).

### Human inter-assessor agreement

ROBoto2 includes trials assessed by more than one person: 52 trials with both an unassisted and an LLM-assisted assessment, and 10 trials with several LLM-assisted assessments (60 assessor pairs). For the 52 trials, the unassisted assessment is the benchmark reference. We used agreement within these pairs as the reference point for each system’s agreement; these values are descriptive and carry no confidence interval. In a post hoc analysis not specified in the protocol, we dichotomized domain and overall judgements as high versus not high. On the 52 trials with both an unassisted and an LLM-assisted assessment, we calculated the sensitivity and specificity of the assisted assessor’s judgements, taking the unassisted assessor’s judgements as the reference, and then the same measures for each AI system against that unassisted assessor. We also calculated sensitivity and specificity for a high-risk overall judgement for every AI system against the reference on the 419 pooled rows. Confidence intervals used the bootstrap described under Statistical analysis.

### Outcomes

The primary outcomes were percentage agreement and Cohen’s κ (21, 24, 25) between each system and the reference at three levels: signalling question (five categories, ROBoto2 trials, unanswered questions scored as no information per benchmark convention), domain (low, some concerns, high) and overall judgement (low, some concerns, high). Secondary outcomes were error direction (overall judgements below the reference category, sensitivity and specificity of high-risk domain judgements against the reference, and domains called high that the reference did not), output validity, the proportion of reader quotations located verbatim in the article (22, 23), and cost and time per trial.

### Statistical analysis

We scored a row without a valid set of signalling answers (due to output failure, parsing failure, or incompleteness) as disagreement at every level. Its predicted class was an additional category, invalid, which never occurs in the reference, reducing agreement and adding a column to the κ confusion matrix with zero chance agreement. A frontier output was valid when all 22 answer lines parsed; a reader output was valid when it completed a parsable record under the grammar.

We derived confidence intervals from 2,000 cluster-bootstrap resamples using the trial as the unit (26, 27), seed 20261103, and percentile 95% intervals. Within each population and cohort, we scored every AI system on the same resampled trials, so paired differences (RoB-BERT minus comparator) use identical resamples. We excluded replicates with undefined κ from that interval and reported their count. Intervals are pointwise and unadjusted for multiplicity; we performed no hypothesis tests, pre-specified no non-inferiority margin, and interpreted intervals directly (28). Component analyses compared the combined system with the reader alone and with the encoder alone on the same resamples.

We pre-specified four sensitivity analyses: SA1, rows where every AI system returned a valid output; SA2, all 1,008 outcome rows; SA3, articles where reader preprocessing truncated input versus those it did not; and SA4, the Cochrane cohort excluding the RECOVERY platform trial comparisons (NCT04381936), which affected one primary row. For descriptive context only, we report the agreement of GPT-4 (2023) on the same Cochrane rows, noting that GPT-4 received the PDF, registration and protocol rather than text alone (6). Hosted-model costs are OpenRouter billings and times are per-request wall time at eight concurrent requests; reader time and cost are batch generation on one H200 accelerator at US$5.00 per hour, excluding model loading; the encoder runs on a workstation CPU at no accelerator cost. The analysis code reproduced the benchmark scorer’s agreement, κ and error-direction counts exactly, and all code, prompts, request records and prediction files are released with the benchmark.

### Post hoc adjudication of disagreements

To determine whether disagreements reflected model error, reference error or defensible differences in judgement, two reviewers (TP and DZ) adjudicated a sample of the 672 domain-level disagreements (341 rows) in the 419 pooled rows: all 26 Cochrane rows (34 domains) and random samples of 40 ROBoto2 rows (73 domains) and 40 LogiMed rows (81 domains), 106 rows and 188 domains in all. Working from written RoB 2 guidance and blinded to provenance, the reviewers received case packets containing the trial text, outcome and both structured assessments (judgements, signalling answers, quotations and rationales) with origin-disclosing phrases removed, and treated registries and protocols as unavailable. The reviewers divided the sample between them and worked independently; each recorded their own judgement before unblinding, then classified the disagreement as RoB-BERT correct, reference correct, both defensible, both wrong or indeterminate, coding error type and severity. Counts pool both reviewers’ judgements; there was no consensus step and no overlap from which to measure inter-reviewer agreement. For benchmark Cochrane trials also assessed in other Cochrane reviews, we verified trial identity by DOI, PMID or title and compared judgements across reviews.

## Results

### Primary outcomes

Pairs of human assessors who rated the same ROBoto2 trial agreed with each other on 42% to 46% of signalling answers, 61% to 65% of domain judgements and 46% to 50% of overall judgements, with κ no higher than 0.35 at any level (Figure 2). On the 185 ROBoto2 trials, RoB-BERT’s domain and overall agreement and κ lay within or above this range, and its signalling-question agreement lay just below it. Claude Fable 5.1 and Gemini 4 Argon lay within the human range at the domain level and for overall agreement but below it for signalling-question agreement and overall κ, and GPT-6 Astra lay below the human range for agreement at every level.

**Figure 2.**
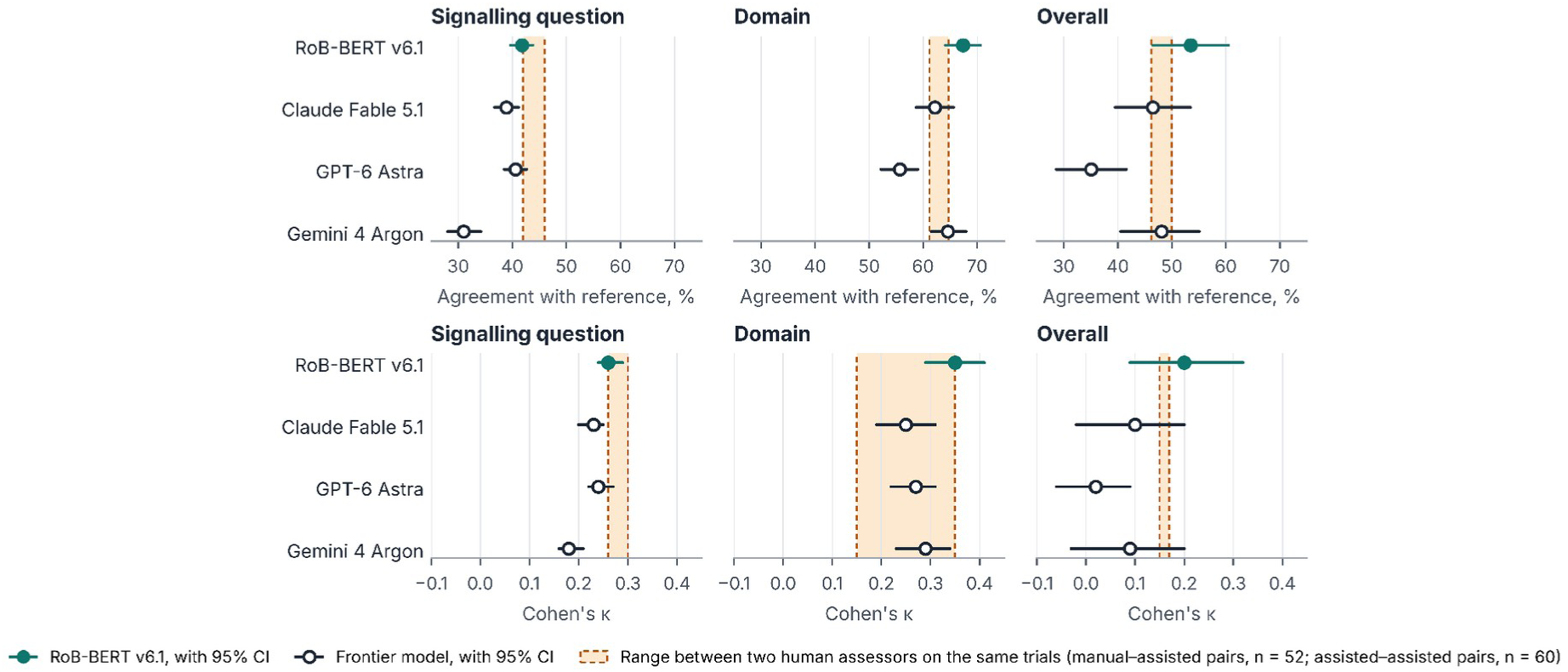
Agreement with the reference compared with agreement between human assessors, ROBoto2 unassisted trials. Percentage agreement (top row) and Cohen’s κ (bottom row) of RoB-BERT v6.1 and each frontier model against the single-assessor reference on the 185 ROBoto2 unassisted trials, with 95% percentile intervals from 2,000 trial-level cluster-bootstrap resamples. The shaded band spans the two inter-assessor values published with the benchmark for ROBoto2 trials assessed by more than one person: an unassisted assessor against an LLM-assisted assessor (52 pairs on 52 trials) and two LLM-assisted assessors (60 pairs on 10 trials). Human values are descriptive and carry no interval; the values are tabulated in Supplementary Table S12.

Table 3 and Figure 2 give agreement and κ for every AI system, and Figure 3 gives the paired differences between RoB-BERT and each frontier model; cohort-level values and paired differences are in Supplementary Tables S4 and S5. At the signalling-question level on the 185 ROBoto2 trials, RoB-BERT, Claude Fable 5.1 and GPT-6 Astra each agreed with the reference on about 40% of answers (κ about 0.25), and Gemini 4 Argon on about 30% (κ below 0.20). RoB-BERT’s agreement was about 3 percentage points higher than Claude Fable 5.1’s and 11 points higher than Gemini 4 Argon’s, with confidence intervals excluding zero, and similar to GPT-6 Astra’s.

**Figure 3.**
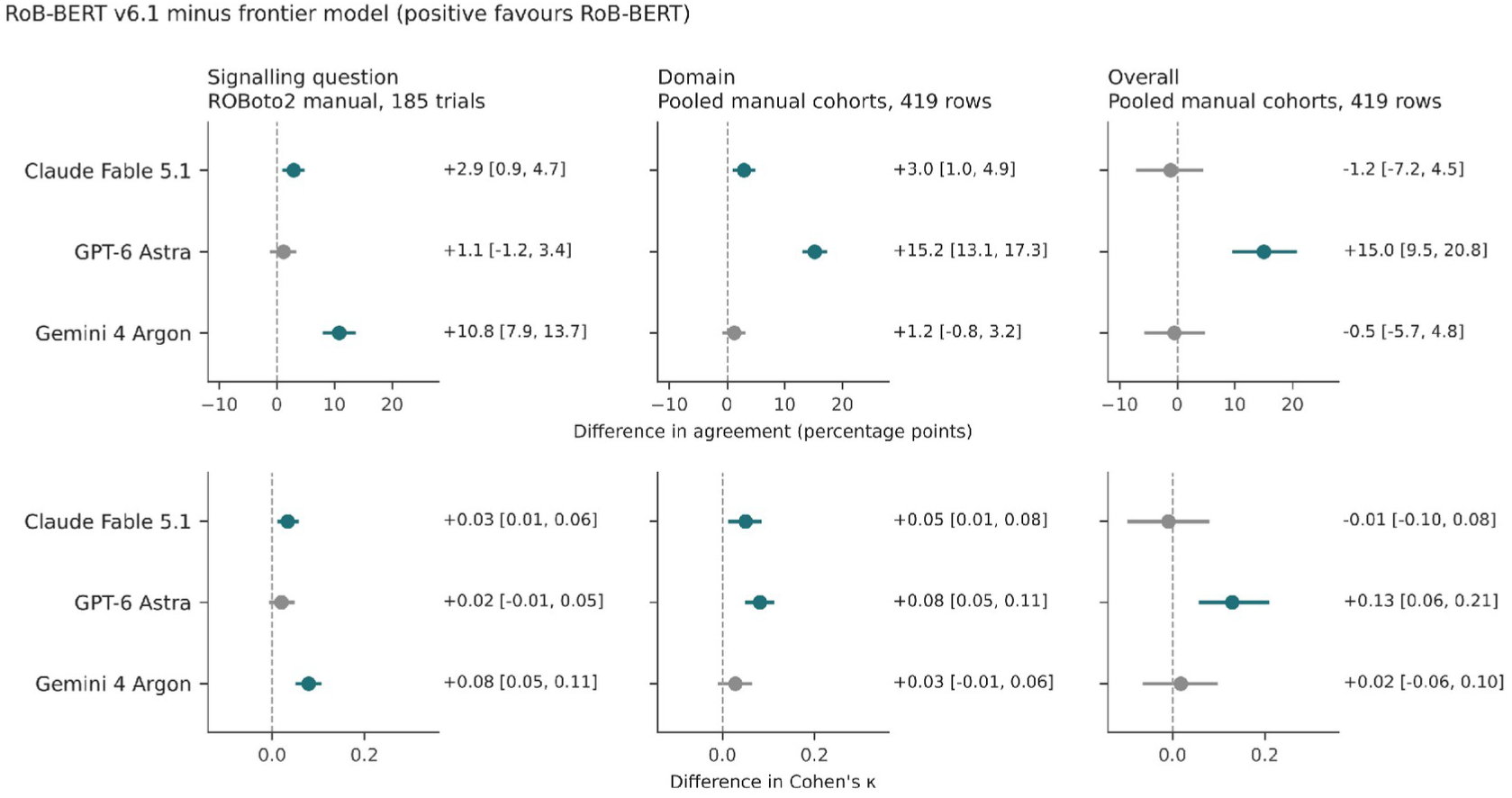
Paired differences between RoB-BERT v6.1 and each frontier model. Difference in percentage agreement (top row) and Cohen’s κ (bottom row), RoB-BERT minus comparator, with 95% percentile intervals from 2,000 trial-level cluster-bootstrap resamples on identical resamples. Signalling-question differences use the 185 ROBoto2 unassisted trials, the only cohort with signalling-level reference answers; domain and overall differences use the 419 pooled rows. Teal, interval excludes zero in favour of RoB-BERT; grey, interval includes zero. Invalid outputs are scored as disagreement.

**Table 3.** Agreement with human reference assessments in the primary population.

| AI system | Valid/rows (ROBoto2 unassisted; pooled) | Signalling question agreement, % (95% CI) | Signalling question κ (95% CI) | Domain agreement, % (95% CI) | Domain κ (95% CI) | Overall agreement, % (95% CI) | Overall κ (95% CI) |
|---|---|---|---|---|---|---|---|
| RoB-BERT v6.1 | 185/185; 419/419 | 41.8 (39.7 to 43.8) | 0.26 (0.24 to 0.29) | 67.9 (65.7 to 70.0) | 0.29 (0.25 to 0.33) | 46.1 (41.3 to 51.1) | 0.16 (0.09 to 0.23) |
| RoB-BERT reader alone | 185/185; 419/419 | 40.8 (38.5 to 43.0) | 0.25 (0.23 to 0.28) | 68.2 (65.9 to 70.3) | 0.29 (0.25 to 0.33) | 46.5 (41.7 to 51.4) | 0.17 (0.10 to 0.24) |
| RoB-BERT encoder alone | 185/185; 419/419 | 42.6 (40.8 to 44.3) | 0.26 (0.24 to 0.28) | 67.0 (65.0 to 69.1) | 0.27 (0.23 to 0.31) | 44.4 (39.5 to 49.2) | 0.05 (-0.02 to 0.13) |
| Claude Fable 5.1 | 185/185; 416/419 | 38.9 (36.7 to 41.2) | 0.23 (0.20 to 0.25) | 65.0 (62.5 to 67.4) | 0.24 (0.20 to 0.28) | 47.3 (42.5 to 52.0) | 0.17 (0.09 to 0.24) |
| GPT-6 Astra | 185/185; 419/419 | 40.6 (38.5 to 42.7) | 0.24 (0.22 to 0.27) | 52.7 (50.7 to 54.9) | 0.21 (0.18 to 0.24) | 31.0 (26.4 to 35.4) | 0.03 (-0.02 to 0.08) |
| Gemini 4 Argon | 184/185; 413/419 | 31.0 (28.0 to 34.2) | 0.18 (0.16 to 0.21) | 66.7 (64.3 to 68.9) | 0.26 (0.22 to 0.30) | 46.5 (41.9 to 51.4) | 0.14 (0.06 to 0.22) |

Signalling-question outcomes use the 185 ROBoto2 trials (22 questions each, five answer categories; an unanswered question is scored as no information). Domain (three categories, five domains per row) and overall (three categories) outcomes use the 419 pooled rows from 418 trials. Intervals are percentile 95% intervals from 2,000 trial-level cluster-bootstrap resamples. Invalid outputs are scored as disagreement at every level. Paired differences are shown in Figure 3 and Supplementary Table S5.

At the domain level across the 419 pooled rows, RoB-BERT, Claude Fable 5.1 and Gemini 4 Argon each agreed with the reference on about two-thirds of judgements (κ 0.24 to 0.29) and GPT-6 Astra on about half (κ 0.21). RoB-BERT’s agreement was about 3 points higher than Claude Fable 5.1’s and 15 points higher than GPT-6 Astra’s, with confidence intervals excluding zero, and similar to Gemini 4 Argon’s.

At the overall level, RoB-BERT, Claude Fable 5.1 and Gemini 4 Argon each agreed with the reference on just under half of judgements (κ 0.14 to 0.17) and GPT-6 Astra on about 30% (κ 0.03). RoB-BERT’s agreement was about 15 points higher than GPT-6 Astra’s and similar to that of the other two. On the 185 ROBoto2 trials alone, RoB-BERT’s overall agreement was 5 to 7 points higher than that of Claude Fable 5.1 and Gemini 4 Argon, with confidence intervals including zero, and 18 points higher than GPT-6 Astra’s.

### Results by cohort

The ranking was not uniform across cohorts (Supplementary Tables S4 to S6). On the Cochrane rows, RoB-BERT had the highest domain agreement of any AI system (84%, against 81% for Claude Fable 5.1 and 61% for GPT-6 Astra) and overall agreement similar to that of Claude Fable 5.1 and Gemini 4 Argon. On the LogiMed rows, domain agreement was about 65% for RoB-BERT, Claude Fable 5.1 and Gemini 4 Argon and 48% for GPT-6 Astra, but RoB-BERT’s overall agreement was about 8 points lower than Claude Fable 5.1’s (37% against 45%) and 6 points lower than Gemini 4 Argon’s; the truncation analysis below accounts for this deficit. On the 151 LLM-assisted ROBoto2 rows, analyzed separately, RoB-BERT was similar to or above every comparator at every level, with confidence intervals excluding zero only against GPT-6 Astra at the domain (about 11 points) and overall (about 15 points) levels.

### Direction of error

The systems differed more in how they erred than in how often (Table 4, Figure 4). GPT-6 Astra called 396 domains high risk, against 117 in the reference, giving the highest sensitivity for high-risk domains (42%) but the lowest specificity (80%), and it rarely rated a trial more favourably than the reference (5% of overall judgements). RoB-BERT, Claude Fable 5.1 and Gemini 4 Argon erred the other way: their sensitivity was 22% to 25% and specificity 92% to 95%, and each rated 17% to 25% of trials more favourably than the reference. The encoder alone called high risk least often (sensitivity 7%, specificity 96%).

**Figure 4.**
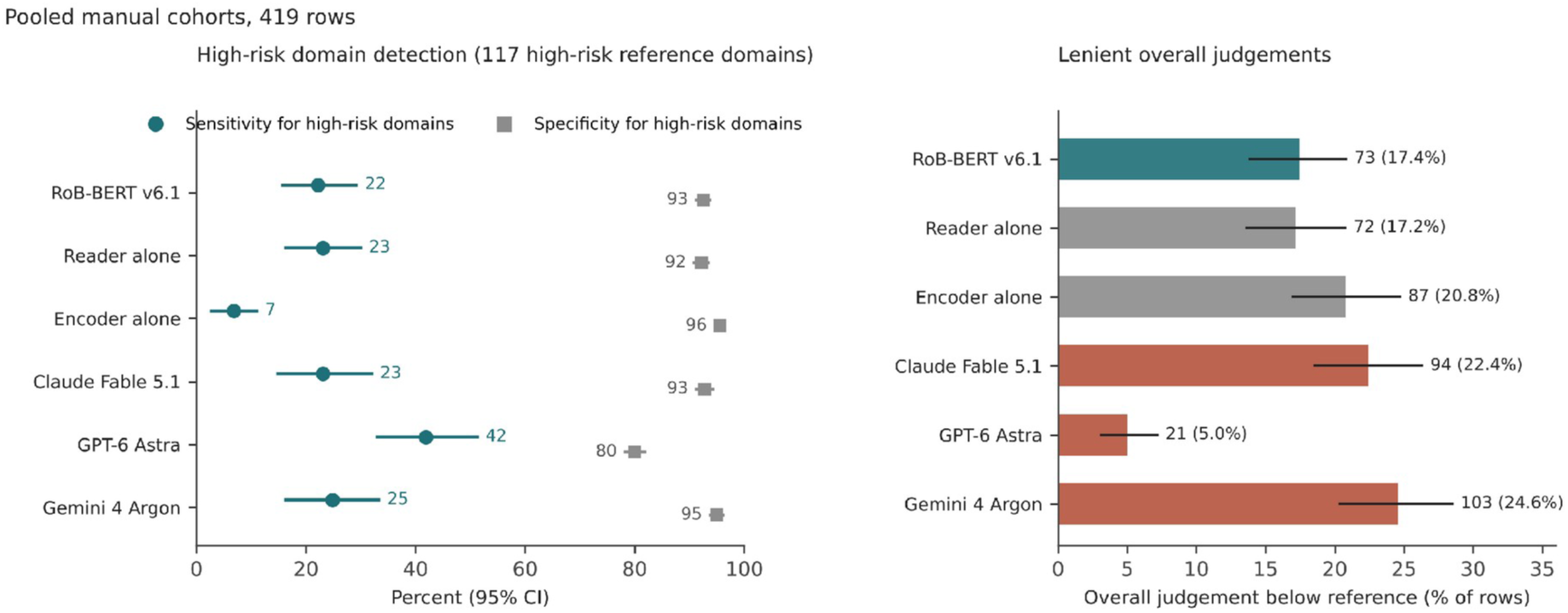
Direction of error on the 419 pooled unassisted rows. Left: sensitivity (circles) and specificity (squares) of high-risk domain judgements against the reference, which contains 117 high-risk domains, with 95% cluster-bootstrap intervals. Right: number and percentage of trials whose overall judgement fell below the reference category (a more favourable rating), with 95% intervals. An invalid output has no judgement and counts as not high.

**Table 4.** Direction of error on the 419 pooled unassisted rows.

| AI system | Overall judgement below the reference category, n (%; 95% CI) | Sensitivity for high-risk domains, % (95% CI) | Specificity for high-risk domains, % (95% CI) | Domains called high that the reference did not, n |
|---|---|---|---|---|
| RoB-BERT v6.1 | 73 (17.4; 13.8 to 20.8) | 22.2 (15.7 to 29.2) | 92.5 (91.2 to 93.7) | 148 |
| RoB-BERT reader alone | 72 (17.2; 13.6 to 20.8) | 23.1 (16.4 to 30.0) | 92.2 (90.9 to 93.5) | 154 |
| RoB-BERT encoder alone | 87 (20.8; 16.9 to 24.7) | 6.8 (2.7 to 11.0) | 95.6 (94.7 to 96.5) | 87 |
| Claude Fable 5.1 | 94 (22.4; 18.5 to 26.3) | 23.1 (14.9 to 32.0) | 92.8 (91.3 to 94.3) | 143 |
| GPT-6 Astra | 21 (5.0; 3.1 to 7.2) | 41.9 (33.0 to 51.3) | 80.0 (78.3 to 81.8) | 396 |
| Gemini 4 Argon | 103 (24.6; 20.3 to 28.5) | 24.8 (16.4 to 33.3) | 95.0 (93.8 to 96.1) | 99 |

The reference contains 117 high-risk domain judgements among 2,095 domain judgements. A judgement below the reference category is a more favourable rating (for example, low where the reference is some concerns). An invalid output has no judgement and counts as not high. Cohort-level values are in Supplementary Table S6.

### High-risk judgements compared with a second human assessor

High-risk domain judgements were the least reproducible between human assessors. On the 52 ROBoto2 trials with both an unassisted and an LLM-assisted assessment, the unassisted (reference) assessor rated 13 domains high risk. Against this reference, the assisted assessor’s sensitivity was 23.1% (3 of 13; 95% CI 0.0 to 50.0) and specificity 90.3% (86.2 to 94.2); between two LLM-assisted assessors, on 10 trials, sensitivity was 33.3% to 36.7% (Supplementary Table S16). On the same 52 trials, RoB-BERT’s sensitivity was 30.8% (4 of 13; 7.7 to 61.5) and specificity 92.7% (89.5 to 95.8); sensitivity and specificity were 23.1% and 91.1% for Claude Fable 5.1, 23.1% and 93.1% for Gemini 4 Argon, and 38.5% and 82.6% for GPT-6 Astra. For the overall judgement on the 419 pooled rows, of which 84 were high risk in the reference, RoB-BERT’s sensitivity was 52.4% (41.6 to 62.9) and specificity 70.1% (65.2 to 75.2), against 39.3% and 77.6% for Claude Fable 5.1, 39.3% and 84.5% for Gemini 4 Argon, and 77.4% and 32.2% for GPT-6 Astra (Supplementary Table S17).

### Benchmark runs and output validity

Every AI system covered all 570 primary and all 1,008 outcome rows. RoB-BERT returned a valid assessment for every primary row and for 1,007 of 1,008 outcome rows (one reader output reached the generation limit). Claude Fable 5.1 returned no answers for 3 of 419 pooled rows (3 of 570 primary rows) and 7 of 1,008 outcome rows, in every case because the provider’s content filter stopped the completion. Gemini 4 Argon returned no or incomplete answers for 6 of 419 pooled rows (8 of 570 primary rows) and 12 of 1,008 outcome rows, in all but one case because it reached the 16,000-token output limit during reasoning. GPT-6 Astra returned a valid output for every row (Supplementary Table S3). Invalid outputs therefore had little effect on agreement: no AI system had more than 6 invalid outputs among the 419 pooled rows, and restricting the analysis to rows with a valid output from every AI system changed estimates by about one point or less (SA1). On the 336 ROBoto2 rows, the protocol runs gave the same signalling answer as the preliminary October runs for 88.2% (Claude Fable 5.1) and 91.8% (GPT-6 Astra) of answers and the same overall judgement for 85.1% and 94.6% of rows (Supplementary Table S2). The reader’s preprocessing truncated input for 250 of 961 requests, almost all LogiMed and Cochrane articles, because the sectioning rules, fixed before the runs, failed to recognize most LogiMed headings; the protocol prohibited changing preprocessing, so we analyzed these rows as run and examined them in SA3.

### Internal evaluation

On the 532 internal test worksheets, the final reader adapter answered signalling questions with κ 0.67 against the audited answers, with domain κ 0.64 and overall κ 0.48, parsed every record, and located 79% of its quotations verbatim in the source text. The selected encoder reached signalling-question κ 0.67, domain κ 0.47 and overall κ 0.31; at the 0.85 threshold it received 48% of answers with 93% accuracy on those answers. On the 16 worksheets assessed independently by one investigator, signalling-question κ was 0.43 for the reader and 0.41 for the encoder.

### Component analyses

The reader alone and the combined system performed almost identically at the domain and overall levels; the encoder gate added +1.0 point (0.1 to 1.9) of signalling-question agreement in the pooled population and nothing beyond it, having replaced 550 of 14,752 reader answers (3.7%). The encoder alone matched the reader at the signalling-question (42.6% versus 40.8%) and domain (67.0% versus 68.2%) levels but carried almost no information about the overall judgement (overall κ 0.05, -0.02 to 0.13, in the pooled population and -0.03 on the ROBoto2 trials) because it defaulted to the majority category where the reader did not; the combined system’s overall κ exceeded the encoder’s by 0.11 (0.02 to 0.20). The discrimination of the overall judgement therefore comes from the generative reader (Supplementary Table S7).

### Sensitivity analyses

Restricting to the 559 primary rows with a valid output from every AI system (SA1) changed estimates by about one point or less and reversed no paired difference (Supplementary Table S8). Extending to all 1,008 outcome rows (SA2) lowered overall agreement for every AI system by up to three points; on the 857 pooled rows, RoB-BERT’s domain agreement was +1.6 points (-0.4 to 3.8) against Claude Fable 5.1, +16.9 (14.0 to 19.8) against GPT-6 Astra and -0.8 (-2.8 to 1.1) against Gemini 4 Argon (Supplementary Table S9). The truncation analysis (SA3) explained the LogiMed pattern: on the 87 pooled rows where preprocessing truncated input, RoB-BERT’s overall agreement was 24.1% (κ -0.14) against 43.7% for Claude Fable 5.1, whereas on the 332 untruncated rows it was 51.8% (κ 0.23) against 48.2% for Claude Fable 5.1 and 48.5% for Gemini 4 Argon, with the highest domain agreement of any AI system (68.4%; +4.1 points, 1.9 to 6.4, against Claude Fable 5.1) (Supplementary Table S10). Removing the RECOVERY comparison (SA4) changed pooled estimates by at most 0.2 points (Supplementary Table S11).

### Evidence grounding, efficiency and descriptive context

The reader located 95.7% (13,472 of 14,082) of its quotations verbatim in the source text (Supplementary Table S3). Hosted frontier models cost US$0.09 to US$0.30 per trial and RoB-BERT US$0.005, 2% to 6% as much; times per trial are in Table 5. For descriptive context, GPT-4 (2023), given the PDF, registration and protocol, agreed with the reference less often on the same 42 Cochrane rows than RoB-BERT did on text alone (Supplementary Table S12, which also tabulates the human inter-assessor values shown in Figure 2).

**Table 5.** Cost, time and output validity on the 570 primary rows.

| AI system | US\$ per trial | US\$ for 570 primary rows | Seconds per trial | Invalid primary rows, n | Configuration |
|---|---|---|---|---|---|
| Claude Fable 5.1 | 0.260 | 148 | 30.4 | 3 | OpenRouter via Anthropic; reasoning effort high; 16,000-token output limit; 8 concurrent requests |
| GPT-6 Astra | 0.301 | 172 | 82.1 | 0 | OpenRouter via OpenAI; reasoning effort high; 16,000-token output limit; 8 concurrent requests |
| Gemini 4 Argon | 0.089 | 51 | 90.8 | 8 | OpenRouter via Google; reasoning effort high; 16,000-token output limit; 8 concurrent requests |
| RoB-BERT reader | 0.005 | 3 | 3.6 | 0 | vLLM 0.31.0 on one H200 at US\$5.00 per hour; batch generation of 961 inputs in 58 minutes; model loading excluded |
| RoB-BERT encoder | 0.000 | 0 | 33.0 | 0 | Production serving script on workstation CPU, 8 threads, 2 concurrent requests; no accelerator |

Hosted-model times are per-request wall time with eight requests in flight; the reader's time is batch throughput (total generation time divided by requests) and the encoder's is CPU serving time, so the three are not directly comparable. Hosted-model costs are OpenRouter billings.

### Post hoc adjudication of disagreements

Of the 188 adjudicated domain-level disagreements, the reviewers judged RoB-BERT correct in 98 (52%), the reference correct in 35 (19%), both defensible in 32 (17%), both wrong in 15 (8%) and 8 (4%) indeterminate (Supplementary Table S13). RoB-BERT was most often judged correct in domain 1 (32 of 46) and domain 5 (31 of 46); in 55 of the 98 domains where the reference was wrong and RoB-BERT correct, the reference had rated domain 1 low risk without information on allocation concealment or domain 5 low risk without an accessible analysis plan. The reference’s errors were mostly misapplied RoB 2 guidance (44, 32 of them major) or leniency on a borderline call (25), and 13 relied on information outside the report, which may reflect reviewer access to a registry or protocol; RoB-BERT’s errors concentrated in domain 3, where 14 of 16 were high-risk calls built on miscounted denominators (Supplementary Table S14). Twenty-three benchmark Cochrane trials had also been assessed by a second, independent Cochrane review team, which agreed with the benchmark reference in 27% to 68% of judgements depending on the domain (Supplementary Table S15).

## Discussion

### Principal findings

Agreement of humans using Cochrane RoB 2 with one another was very poor, with κ uniformly less than 0.4 and often appreciably lower. A 27-billion-parameter reader fine-tuned on audited RoB 2 worksheets, paired with a biomedical encoder and the published decision algorithm, showed agreement similar to that between humans, matched or exceeded three frontier LLMs on identical trial text, and did so at 2% to 6% of their cost. RoB-BERT agreed with the reference more often than Claude Fable 5.1 and Gemini 4 Argon at the signalling-question level, more often than Claude Fable 5.1 and GPT-6 Astra at the domain level, and much more often than GPT-6 Astra at the overall level; otherwise it performed similarly to the frontier models. It returned a valid assessment for every primary row, and almost all of its quotations could be found verbatim in the source text. On articles the reader saw in full, it had the highest domain-level agreement of any AI system. Like Claude Fable 5.1 and Gemini 4 Argon, RoB-BERT identified fewer than a quarter of the domains the reference rated high risk, about as many as a second human assessor identified on the same trials.

### Interpretation

The skewed distribution of judgements, with low risk predominating in most domains, may have contributed to the low κ values (24, 25). Even absolute agreement was limited, however: human assessors agreed with one another on less than 65% of domain judgements and on 50% or less of overall judgements. Agreement between human assessors is the appropriate yardstick for judging the performance of the AI systems, and the comparison is therefore with a flawed instrument. Low reliability is not inevitable in risk-of-bias assessment: junior researchers using the more recently developed ROBUST-RCT tool (29) achieved at least moderate agreement (30). In our study, RoB-BERT’s agreement with the reference at the domain and overall levels was similar to the agreement between humans, and the best frontier models achieved this at the domain level and for overall percentage agreement but not for overall κ.

Adjudication suggests that RoB-BERT’s disagreements with the reference often reflected strict application of the RoB 2 rules rather than error, although the adjudicators included the system’s developer. The commonest pattern was a reference rating of low risk for randomization or for selection of the reported result where the report gave no information on allocation concealment or no accessible analysis plan, which may reflect pragmatic leniency or unrecorded access to a registry or protocol; RoB-BERT, which saw only the report, applied the written rules. Its own characteristic error, a high-risk call on a miscounted denominator in domain 3, is visible in the quoted evidence and can be targeted in training. One in six disagreements was judged defensible on both sides, a component of RoB 2 agreement that no system can remove.

The component analyses separate the roles of the two parts. The encoder is a high-confidence gate: where it is certain it overrides the reader and slightly improves signalling-question agreement, but where it is uncertain it defaults to the majority answer, low risk. Because the overall RoB 2 judgement is set by the worst domain, a model that defaults to low risk when uncertain has an overall κ near zero. The generative reader must commit to an answer for every question and support it with quoted text, and it supplies nearly all of the signal for the overall judgement.

The truncation finding is a limitation of this version of the system, not of the comparison, and is fixable. The frontier models received the whole article; the reader saw a shortened, section-prioritized window because the fixed sectioning rules failed on the heading conventions of LogiMed’s source journals, losing the outcome-reporting and analysis-plan material that drives the domain 3 to 5 judgements. Correcting the sectioning rules or extending the context would be the first change in a subsequent version, and SA3 shows the expected gain.

### Comparison with previous work

Earlier evaluations of general-purpose chatbots for RoB 2 reported agreement in the range of the frontier models here. GPT-4 on Cochrane assessments reached domain κ near zero despite access to protocols and registrations (6); ChatGPT-4o gave moderate domain agreement with substantial variation across domains (7); and an LLM-assisted workflow improved speed more than agreement (8). ROBoto2 found that showing assessors an LLM’s suggestions changed their answers, which is why the assisted cohort is reported separately (5). LogiMed-RoB showed that LLMs often give domain judgements inconsistent with their own signalling answers (13); passing every AI system’s answers through the same algorithm removes that failure mode here.

Encoder-only models for evidence quality assessment have been limited by the lack of grounding (12); the present system gives the quotation task to the reader and the confidence task to the encoder. No previous study evaluated a fine-tuned RoB 2 system and current frontier models on identical text with recorded settings and a pre-registered analysis.

### Strengths and limitations

The evaluation was registered prospectively under an unamended protocol, with benchmark texts, reference assessments, model weights and prompts frozen and checksummed. The development corpus was screened against the benchmark by trial identifier, DOI and title so that no benchmark trial was used in training. Every AI system received identical text and invalid outputs were scored as disagreement rather than excluded. Intervals were clustered by trial on paired resamples and all prediction files, prompts, request records and analysis code are public. The study has limitations. First, each reference is a single published judgement, made by one assessor or agreed by two, rather than an expert reference standard, so it carries the limited inter-rater reliability of RoB 2. Second, preprocessing truncated the text of a subset of articles, mostly in the LogiMed cohort, removing context for domain judgements in those trials. Third, frontier-model performance is sensitive to prompt and decoding settings, and we tested one standardized configuration. Fourth, the adjudication was post hoc, by two reviewers who are authors of this study, on a sample of disagreements divided between them, with no consensus step and no measure of inter-reviewer agreement, and its findings are descriptive. Fifth, we assembled the benchmark as well as the system under test; the reference assessments come from three independently published sources and were frozen before any run, but the choice of sources and scoring conventions was ours. Sixth, every pair of human assessors used as the inter-assessor reference included at least one assessor who had seen LLM suggestions, which may have increased their agreement with each other. Given how poor that agreement was, and because any such inflation would make the human yardstick harder rather than easier to meet, this is unlikely to be a major problem. Seventh, the comparison of high-risk judgements was post hoc and rests on 52 trials with 13 high-risk reference domains, so its confidence intervals are wide. Finally, we measured offline benchmark performance on report text, not reviewer workflow efficiency or usability in live reviews.

### Implications

For evidence synthesis teams, these findings support RoB-BERT as a first-pass assessor whose output a reviewer – or given the poor agreement among human reviewers, probably two reviewers – must then check. At half a cent per trial with deterministic, complete output, it makes assessing every trial at every update of a living review affordable, which hosted models at many times the cost do not. Because almost all of its quotations can be located in the report, a reviewer can audit each answer against the quoted sentence rather than re-read the trial.

## Conclusion

Human agreement on RoB 2 was poor, with κ values uniformly less than 0.4. A task-specific fine-tuned system matched or exceeded three frontier LLMs in agreement with human RoB 2 assessments at the signalling-question and domain levels, matched the best of them at the overall level, and did so at a small fraction of the cost with complete output in which each answer cites the supporting text. Its domain and overall agreement with single human assessors was within or above the agreement between two human assessors rating the same trial, it identified the reference’s high-risk domains as often as a second assessor did, and every system tended to err in a direction that matters for certainty ratings. Fine-tuned systems are a credible basis for assisted assessment, potentially as one rater in a consensus process; they are not a replacement for the reviewer.

## Data Availability

The RoB 2 AI Benchmark v1.0.0, including labels, texts where licensing permits, the scorer and the RoB 2 algorithm, is available at https://github.com/CoreSR/rob2-ai-benchmark. Prompts, request records, prediction files, the methods log and the analysis code are released with the results.

## Declarations

### Contributors

TP conceived the study, built the system, wrote the protocol, ran the evaluation and analysis, and drafted the manuscript. JM, AAz, DN, DZ, AAg, and GG contributed to the study design and interpretation of results and revised the manuscript critically. All authors approved the final version. TP is the guarantor.

### Ethics

The study used published trial reports and previously published assessments and involved no human participants; ethics approval was not required.

### Data and code availability

The RoB 2 AI Benchmark v1.0.0, including labels, texts where licensing permits, the scorer and the RoB 2 algorithm, is available at github.com/CoreSR/rob2-ai-benchmark. Prompts, request records, prediction files, the methods log and the analysis code are released with the results.

## References

1. Guyatt G, Wang Y, Eachempati P, Iorio A, Murad MH, Hultcrantz M, et al. Core GRADE 4: rating certainty of evidence: risk of bias, publication bias, and reasons for rating up certainty. BMJ. 2025;389:e083864.

2. Sterne JAC, Savović J, Page MJ, Elbers RG, Blencowe NS, Boutron I, et al. RoB 2: a revised tool for assessing risk of bias in randomised trials. BMJ. 2019;366:l4898.

3. Crocker TF, Lam N, Jordão M, Brundle C, Prescott M, Forster A, et al. Risk-of-bias assessment using Cochrane’s revised tool for randomized trials (RoB 2) was useful but challenging and resource-intensive: observations from a systematic review. J Clin Epidemiol. 2023;161:39–45.

4. Minozzi S, Dwan K, Borrelli F, Filippini G. Reliability of the revised Cochrane risk-of-bias tool for randomised trials (RoB2) improved with the use of implementation instruction. J Clin Epidemiol. 2022;141:99–105.

5. Hevia A, Chintalapati S, Lai VKW, Tam NT, Wong W-T, Klassen TP, et al. ROBOTO2: an interactive system and dataset for LLM-assisted clinical trial risk of bias assessment. In: Proceedings of the 2025 Conference on Empirical Methods in Natural Language Processing: System Demonstrations. Suzhou, China: Association for Computational Linguistics; 2025. p. 12-25.

6. Pitre T, Jassal T, Talukdar JR, Shahab M, Ling M, Zeraatkar D. ChatGPT for assessing risk of bias of randomized trials using the RoB 2.0 tool: a methods study. medRxiv. 2026:2023.11.19.23298727.

7. Rose CJ, Bidonde J, Ringsten M, Glanville J, Potrebny T, Cooper C, et al. Using a large language model (ChatGPT-4o) to assess the risk of bias in randomized controlled trials of medical interventions: interrater agreement with human reviewers. Cochrane Evid Synth Methods. 2025;3(5):e70048.

8. Huang J, Lai H, Zhao W, Xia D, Bai C, Sun M, et al. Large language model-assisted risk-of-bias assessment in randomized controlled trials using the revised risk-of-bias tool: evaluation study. J Med Internet Res. 2025;27:e70450.

9. Devlin J, Chang M-W, Lee K, Toutanova K. BERT: pre-training of deep bidirectional transformers for language understanding. In: Proceedings of the 2019 Conference of the North American Chapter of the Association for Computational Linguistics: Human Language Technologies, Volume 1. Minneapolis, MN: Association for Computational Linguistics; 2019. p. 4171-86.

10. Alsentzer E, Murphy J, Boag W, Weng W-H, Jindi D, Naumann T, et al. Publicly available clinical BERT embeddings. In: Proceedings of the 2nd Clinical Natural Language Processing Workshop. Minneapolis, MN: Association for Computational Linguistics; 2019. p. 72-8.

11. Yasunaga M, Leskovec J, Liang P. LinkBERT: pretraining language models with document links. In: Proceedings of the 60th Annual Meeting of the Association for Computational Linguistics, Volume 1. Dublin, Ireland: Association for Computational Linguistics; 2022. p. 8003-16.

12. Šuster S, Baldwin T, Lau JH, Jimeno Yepes A, Martinez Iraola D, Otmakhova Y, et al. Automating quality assessment of medical evidence in systematic reviews: model development and validation study. J Med Internet Res. 2023;25:e35568.

13. Huang J, Tang Z, Ling Q, Kuang Z, E H. Can LLMs follow medical expert logic? A benchmark for hierarchical logical consistency in risk-of-bias assessment. arXiv; 2026.

14. Sounack T, Davis J, Durieux B, Chaffin A, Pollard TJ, Lehman E, et al. BioClinical ModernBERT: a state-of-the-art long-context encoder for biomedical and clinical NLP. arXiv; 2025.

15. Guo C, Pleiss G, Sun Y, Weinberger KQ. On calibration of modern neural networks. In: Precup D, Teh YW, editors. Proceedings of the 34th International Conference on Machine Learning. Proceedings of Machine Learning Research, vol 70. PMLR; 2017. p. 1321-30.

16. Gallifant J, Afshar M, Ameen S, Aphinyanaphongs Y, Chen S, Cacciamani G, et al. The TRIPOD-LLM reporting guideline for studies using large language models. Nat Med. 2025;31(1):60–9.

17. Dettmers T, Pagnoni A, Holtzman A, Zettlemoyer L. QLoRA: efficient finetuning of quantized LLMs. arXiv; 2023.

18. Dao T, Fu DY, Ermon S, Rudra A, Ré C. FlashAttention: fast and memory-efficient exact attention with IO-awareness. arXiv; 2022.

19. Chen T, Xu B, Zhang C, Guestrin C. Training deep nets with sublinear memory cost. arXiv; 2016.

20. OpenRouter Inc. OpenRouter API reference and parameters. 2026.

21. Cohen J. A coefficient of agreement for nominal scales. Educ Psychol Meas. 1960;20(1):37–46.

22. Kryscinski W, McCann B, Xiong C, Socher R. Evaluating the factual consistency of abstractive text summarization. In: Proceedings of the 2020 Conference on Empirical Methods in Natural Language Processing (EMNLP). Online: Association for Computational Linguistics; 2020. p. 9332-46.

23. Laban P, Schnabel T, Bennett PN, Hearst MA. SummaC: re-visiting NLI-based models for inconsistency detection in summarization. Trans Assoc Comput Linguist. 2022;10:163–77.

24. Feinstein AR, Cicchetti DV. High agreement but low kappa: I. The problems of two paradoxes. J Clin Epidemiol. 1990;43(6):543–9.

25. Byrt T, Bishop J, Carlin JB. Bias, prevalence and kappa. J Clin Epidemiol. 1993;46(5):423–9.

26. Efron B, Tibshirani RJ. An introduction to the bootstrap. 1st ed. Boca Raton, FL: Chapman and Hall/CRC; 1994.

27. Field CA, Welsh AH. Bootstrapping clustered data. J R Stat Soc Series B Stat Methodol. 2007;69(3):369–90.

28. Piaggio G, Elbourne DR, Pocock SJ, Evans SJW, Altman DG, CONSORT Group. Reporting of noninferiority and equivalence randomized trials: extension of the CONSORT 2010 statement. JAMA. 2012;308(24):2594–604.

29. Wang Y, Keitz S, Briel M, Glasziou P, Brignardello-Petersen R, Siemieniuk RAC, et al. Development of ROBUST-RCT: Risk Of Bias instrument for Use in SysTematic reviews-for Randomised Controlled Trials. BMJ. 2025;388:e081199.

30. Vidor PR, Rossi Fermo SS, Casiraghi Y, Moraes de Souza A, Falavigna M, Ziegelmann PK, et al. The ROBUST-RCT tool had at least moderate agreement in an inter-rater reliability study of assessments by junior researchers. J Clin Epidemiol. 2026;198:112418.

