## Supplementary Material for "Task-specific fine-tuning versus frontier large language models for RoB 2 risk of bias assessment: development and external benchmark evaluation of RoB-BERT"

#### Contents

Supplementary Table S1. Scoring record.

Supplementary Table S2. Run-to-run variation of frontier models: preliminary 12 October 2026 runs versus the protocol runs, 336 ROBoto2 rows.

Supplementary Table S3. Output validity on all 1,008 outcome rows.

Supplementary Table S4. Agreement by cohort, primary population.

Supplementary Table S5. Paired differences, RoB-BERT v6.1 minus each frontier model, by cohort.

Supplementary Table S6. Direction of error by cohort, primary population.

Supplementary Table S7. Component analyses: combined system minus reader alone and minus encoder alone.

Supplementary Table S8. Sensitivity analysis SA1: rows with a valid output from every AI system (559 of 570 primary rows).

Supplementary Table S9. Sensitivity analysis SA2: all outcome rows with text (1,008 rows; 857 pooled unassisted rows).

Supplementary Table S10. Sensitivity analysis SA3: articles truncated by the reader's preprocessing versus not truncated, primary population.

Supplementary Table S11. Sensitivity analysis SA4: Cochrane cohort without the RECOVERY platform trial comparison (NCT04381936).

Supplementary Table S12. Descriptive context: human inter-assessor agreement and the earlier GPT-4 evaluation.

Supplementary Table S13. Adjudication of disagreements by domain (unweighted counts, 188 domains).

Supplementary Table S14. Types of error attributed to the reference and to RoB-BERT v6.1 in the adjudicated disagreements.

Supplementary Table S15. Agreement between the benchmark reference and an independent Cochrane review team that assessed the same trial.

Supplementary Table S16. High-risk judgements: sensitivity and specificity between human assessors, and between each system and the same assessors, on ROBoto2 trials rated by more than one assessor.

Supplementary Table S17. Sensitivity and specificity for a high-risk overall judgement, 419 pooled unassisted rows and subgroups.

All intervals are percentile 95% intervals from 2,000 trial-level cluster-bootstrap resamples (seed 20261103); paired differences use identical resamples. Invalid outputs are scored as disagreement at every level. Δ, RoB-BERT v6.1 minus comparator; n/a, not applicable.

### Supplementary Table S1. Scoring record.

| Item | Value |
|---|---|
| Protocol | RoB-BERT Fine-Tuned Models Protocol v1.3, not amended |
| Benchmark | rob2-ai-benchmark v1.0.0, commit 81a3047 |
| Labels SHA-256 | 674957e9996d3c1ee1a54a6c35a7020940edd73a6765861c9cc57867f9fb6417 |
| RoB 2 algorithm | rob2bench/rob2.py at commit 81a3047 (published RoB 2 decision tables) |
| Reader adapter | tpitre3/rob2-hub3-qwen-stage2-sft on Qwen/Qwen3.8-27B; vLLM 0.31.0, Transformers 5.17.0; temperature 0; grammar on; context 16,384; 3,072 new tokens |
| Encoder | tpitre3/rob2-hub3-head-W-s32, tag v6.1.0 (BioClinical ModernBERT-large, 8,192 tokens); gate threshold 0.85 |
| Frontier settings | OpenRouter; reasoning effort high; max_tokens 16,000; provider defaults otherwise; neutral system prompt SHA-256 cccb1f56ec5a7d51388ae0f20fe41972706364272f086cf5a794bc5b19e40c28 |
| Bootstrap | 2,000 trial-level cluster resamples; seed 20261103; percentile 95% intervals; identical resamples within each population and cohort |
| Invalid handling | Invalid or missing row scored as class INVALID at every level (disagreement; extra κ column never present in the reference) |
| Rows | Primary 570; all outcomes 1,008 (protocol text: 1,001); S1 all valid 559 |
| Prediction file: RoB-BERT v6.1 | b3d1180d53c98da15f69ecd7893fa8b839c80bfcbf5c7ad0f53503580274a0f4 |
| Prediction file: RoB-BERT v6.1 reader alone | 2eb6aa0f247122ac85d27f170ec2e810ccc8ee44fb625abc68fe01fe27b25f27 |
| Prediction file: RoB-BERT v6.1 encoder alone (W-s32) | 3c4b4843da6e2ac54cf2358ded1b05f5858e818b5d6ea1f530093421975b3126 |
| Prediction file: Claude Fable 5.1 | 114ccba7cbe0d19616f0b48d08df82aab1e66f04e3e9b7b7a39129d965b39e3c |
| Prediction file: GPT-6 Astra | 2d586fe0c4515eb08db0ca10ea974408765792a3e103a5cbb5725ef55f426931 |
| Prediction file: Gemini 4 Argon | 5d33f0b24d61087b79e7fa37805a877a0e2371f2dec0e100b95fb950659379c1 |

### Supplementary Table S2. Run-to-run variation of frontier models: preliminary 12 October 2026 runs versus the protocol runs, 336 ROBoto2 rows.

| Model | Rows | Same signalling answer, % | Same domain judgement, % | Same overall judgement, % |
|---|---|---|---|---|
| Claude Fable 5.1 | 336 | 88.2 | 90.2 | 85.1 |
| GPT-6 Astra | 336 | 91.8 | 96.1 | 94.6 |

The preliminary October runs used a 32,000-token output limit for GPT-6 Astra and the default reasoning effort for Claude Fable 5.1 and are not used in any analysis. Gemini 4 Argon was not run in October.

**Supplementary Table S3. Output validity on all 1,008 outcome rows.**

| AI system | Valid outputs | Failures |
|---|---|---|
| RoB-BERT v6.1 (reader) | 960 of 961 requests (1,007 of 1,008 rows) | One completion reached the 3,072-token generation limit |
| Claude Fable 5.1 | 1,001 of 1,008 rows | 7 rows with no answers: completion stopped by the provider's content filter |
| GPT-6 Astra | 1,008 of 1,008 rows | None |
| Gemini 4 Argon | 996 of 1,008 rows | 11 rows with no answers (16,000-token output limit reached) and 1 row with 17 of 22 answers |

Reader evidence on the 961 unique requests: 13,472 of 14,082 quotations (95.7%) located verbatim in the article; 250 inputs truncated; the encoder gate replaced 550 of 14,752 reader answers; no output contained reviewer audit text.

**Supplementary Table S4. Agreement by cohort, primary population.**

| AI system | Valid/rows | Signalling question agreement, % (95% CI) | Signalling question κ (95% CI) | Domain agreement, % (95% CI) | Domain κ (95% CI) | Overall agreement, % (95% CI) | Overall κ (95% CI) |
|---|---|---|---|---|---|---|---|
| **ROBoto2 unassisted (185 rows, 185 trials)** | | | | | | | |
| RoB-BERT v6.1 | 185/185 | 41.8 (39.7 to 43.8) | 0.26 (0.24 to 0.29) | 67.4 (64.1 to 70.6) | 0.35 (0.29 to 0.41) | 53.5 (46.5 to 60.5) | 0.20 (0.09 to 0.32) |
| RoB-BERT reader alone | 185/185 | 40.8 (38.5 to 43.0) | 0.25 (0.23 to 0.28) | 67.6 (64.2 to 70.9) | 0.35 (0.29 to 0.41) | 53.5 (46.5 to 60.5) | 0.21 (0.10 to 0.32) |
| RoB-BERT encoder alone | 185/185 | 42.6 (40.8 to 44.3) | 0.26 (0.24 to 0.28) | 68.2 (65.3 to 71.1) | 0.36 (0.31 to 0.41) | 47.0 (40.0 to 54.6) | -0.03 (-0.14 to 0.09) |
| Claude Fable 5.1 | 185/185 | 38.9 (36.7 to 41.2) | 0.23 (0.20 to 0.25) | 62.2 (58.7 to 65.7) | 0.25 (0.19 to 0.31) | 46.5 (39.5 to 53.5) | 0.10 (-0.02 to 0.20) |
| GPT-6 Astra | 185/185 | 40.6 (38.5 to 42.7) | 0.24 (0.22 to 0.27) | 55.7 (52.2 to 59.0) | 0.27 (0.22 to 0.31) | 35.1 (28.6 to 41.6) | 0.02 (-0.06 to 0.09) |
| Gemini 4 Argon | 184/185 | 31.0 (28.0 to 34.2) | 0.18 (0.16 to 0.21) | 64.6 (61.5 to 68.0) | 0.29 (0.23 to 0.34) | 48.1 (40.5 to 55.1) | 0.09 (-0.03 to 0.20) |
| **LogiMed-RoB (192 rows, 192 trials)** | | | | | | | |
| RoB-BERT v6.1 | 192/192 | n/a | n/a | 65.0 (61.7 to 68.5) | 0.16 (0.10 to 0.22) | 36.5 (30.2 to 43.2) | 0.04 (-0.06 to 0.14) |
| RoB-BERT reader alone | 192/192 | n/a | n/a | 65.6 (62.3 to 69.1) | 0.16 (0.11 to 0.22) | 37.5 (30.7 to 44.3) | 0.06 (-0.04 to 0.15) |
| RoB-BERT encoder alone | 192/192 | n/a | n/a | 62.4 (59.4 to 65.4) | 0.12 (0.07 to 0.17) | 38.0 (31.2 to 45.3) | 0.03 (-0.07 to 0.12) |
| Claude Fable 5.1 | 189/192 | n/a | n/a | 64.2 (60.1 to 68.1) | 0.18 (0.11 to 0.24) | 44.8 (37.5 to 52.1) | 0.16 (0.05 to 0.26) |
| GPT-6 Astra | 192/192 | n/a | n/a | 48.2 (45.2 to 51.1) | 0.13 (0.09 to 0.16) | 29.2 (22.9 to 35.9) | 0.03 (-0.04 to 0.09) |
| Gemini 4 Argon | 189/192 | n/a | n/a | 65.7 (62.3 to 69.3) | 0.18 (0.12 to 0.24) | 42.2 (35.4 to 49.0) | 0.11 (0.01 to 0.21) |
| **Cochrane (42 rows, 41 trials)** | | | | | | | |

| AI system | Valid/rows | Signalling question agreement, % (95% CI) | Signalling question κ (95% CI) | Domain agreement, % (95% CI) | Domain κ (95% CI) | Overall agreement, % (95% CI) | Overall κ (95% CI) |
|---|---|---|---|---|---|---|---|
| RoB-BERT v6.1 | 42/42 | n/a | n/a | 83.8 (79.0 to 88.6) | 0.49 (0.36 to 0.61) | 57.1 (42.8 to 73.2) | 0.34 (0.12 to 0.56) |
| RoB-BERT reader alone | 42/42 | n/a | n/a | 82.9 (77.6 to 87.9) | 0.45 (0.33 to 0.57) | 57.1 (42.8 to 73.2) | 0.34 (0.12 to 0.56) |
| RoB-BERT encoder alone | 42/42 | n/a | n/a | 82.9 (78.0 to 87.4) | 0.41 (0.28 to 0.51) | 61.9 (46.3 to 76.1) | 0.33 (0.10 to 0.55) |
| Claude Fable 5.1 | 42/42 | n/a | n/a | 81.0 (74.9 to 86.5) | 0.42 (0.28 to 0.55) | 61.9 (46.5 to 76.2) | 0.36 (0.13 to 0.59) |
| GPT-6 Astra | 42/42 | n/a | n/a | 60.5 (54.4 to 66.8) | 0.28 (0.18 to 0.38) | 21.4 (9.5 to 34.1) | 0.02 (-0.09 to 0.13) |
| Gemini 4 Argon | 40/42 | n/a | n/a | 80.0 (71.7 to 87.3) | 0.38 (0.21 to 0.55) | 59.5 (44.2 to 74.4) | 0.34 (0.08 to 0.58) |
| **Pooled unassisted cohorts (419 rows, 418 trials)** | | | | | | | |
| RoB-BERT v6.1 | 419/419 | 41.8 (39.8 to 43.7) | 0.26 (0.24 to 0.28) | 67.9 (65.7 to 70.0) | 0.29 (0.25 to 0.33) | 46.1 (41.3 to 51.1) | 0.16 (0.09 to 0.23) |
| RoB-BERT reader alone | 419/419 | 40.8 (38.6 to 42.9) | 0.25 (0.23 to 0.28) | 68.2 (65.9 to 70.3) | 0.29 (0.25 to 0.33) | 46.5 (41.7 to 51.4) | 0.17 (0.10 to 0.24) |
| RoB-BERT encoder alone | 419/419 | 42.6 (40.9 to 44.4) | 0.26 (0.24 to 0.28) | 67.0 (65.0 to 69.1) | 0.27 (0.23 to 0.31) | 44.4 (39.5 to 49.2) | 0.05 (-0.02 to 0.13) |
| Claude Fable 5.1 | 416/419 | 38.9 (36.8 to 41.0) | 0.23 (0.20 to 0.25) | 65.0 (62.5 to 67.4) | 0.24 (0.20 to 0.28) | 47.3 (42.5 to 52.0) | 0.17 (0.09 to 0.24) |
| GPT-6 Astra | 419/419 | 40.6 (38.5 to 42.8) | 0.24 (0.22 to 0.27) | 52.7 (50.7 to 54.9) | 0.21 (0.18 to 0.24) | 31.0 (26.4 to 35.4) | 0.03 (-0.02 to 0.08) |
| Gemini 4 Argon | 413/419 | 31.0 (27.8 to 34.1) | 0.18 (0.15 to 0.21) | 66.7 (64.3 to 68.9) | 0.26 (0.22 to 0.30) | 46.5 (41.9 to 51.4) | 0.14 (0.06 to 0.22) |
| **ROBoto2 assisted (151 rows, 151 trials)** | | | | | | | |
| RoB-BERT v6.1 | 151/151 | 37.5 (35.3 to 39.7) | 0.22 (0.19 to 0.24) | 62.9 (59.2 to 66.6) | 0.27 (0.22 to 0.33) | 53.0 (45.0 to 60.9) | 0.19 (0.05 to 0.33) |
| RoB-BERT reader alone | 151/151 | 37.9 (35.5 to 40.2) | 0.22 (0.19 to 0.25) | 62.6 (59.1 to 66.4) | 0.27 (0.21 to 0.33) | 52.3 (44.4 to 60.3) | 0.19 (0.04 to 0.33) |
| RoB-BERT encoder alone | 151/151 | 37.6 (35.6 to 39.7) | 0.22 (0.19 to 0.24) | 63.0 (59.9 to 66.2) | 0.26 (0.21 to 0.32) | 51.0 (43.0 to 58.9) | 0.02 (-0.08 to 0.13) |
| Claude Fable 5.1 | 151/151 | 36.2 (34.1 to 38.2) | 0.19 (0.17 to 0.22) | 61.6 (58.4 to 64.8) | 0.25 (0.19 to 0.30) | 47.7 (39.7 to 55.6) | 0.09 (-0.04 to 0.22) |
| GPT-6 Astra | 151/151 | 36.2 (34.4 to 38.0) | 0.21 (0.18 to 0.23) | 52.3 (49.0 to 55.8) | 0.20 (0.15 to 0.25) | 38.4 (31.1 to 46.4) | -0.04 (-0.15 to 0.07) |
| Gemini 4 Argon | 149/151 | 39.8 (37.2 to 42.3) | 0.23 (0.20 to 0.25) | 63.0 (58.9 to 66.9) | 0.28 (0.22 to 0.33) | 49.0 (41.0 to 57.0) | 0.14 (0.01 to 0.27) |

Signalling-question reference answers exist only for ROBoto2; n/a, not applicable. The ROBoto2 assisted cohort's reference was produced with LLM suggestions on screen and is reported separately, never pooled.

**Supplementary Table S5. Paired differences, RoB-BERT v6.1 minus each frontier model, by cohort.**

| Comparison | Signalling question Δ agreement, points (95% CI) | Signalling question Δκ (95% CI) | Domain Δ agreement, points (95% CI) | Domain Δκ (95% CI) | Overall Δ agreement, points (95% CI) | Overall Δκ (95% CI) |
|---|---|---|---|---|---|---|
| **ROBoto2 unassisted (185 rows)** | | | | | | |
| RoB-BERT minus Claude Fable 5.1 | +2.9 (0.9 to 4.7) | +0.03 (0.01 to 0.06) | +5.2 (2.1 to 8.3) | +0.10 (0.05 to 0.16) | +7.0 (-1.6 to 15.7) | +0.11 (-0.03 to 0.25) |
| RoB-BERT minus GPT-6 Astra | +1.1 (-1.2 to 3.4) | +0.02 (-0.01 to 0.05) | +11.7 (8.6 to 14.8) | +0.08 (0.04 to 0.13) | +18.4 (11.4 to 25.9) | +0.19 (0.07 to 0.30) |
| RoB-BERT minus Gemini 4 Argon | +10.8 (7.9 to 13.7) | +0.08 (0.05 to 0.11) | +2.7 (-0.2 to 5.8) | +0.06 (0.01 to 0.12) | +5.4 (-2.2 to 13.5) | +0.12 (0.00 to 0.24) |
| **LogiMed-RoB (192 rows)** | | | | | | |
| RoB-BERT minus Claude Fable 5.1 | n/a | n/a | +0.8 (-2.1 to 3.9) | -0.02 (-0.07 to 0.04) | -8.3 (-16.7 to -0.5) | -0.12 (-0.25 to -0.01) |
| RoB-BERT minus GPT-6 Astra | n/a | n/a | +16.8 (13.6 to 20.0) | +0.03 (-0.02 to 0.08) | +7.3 (-1.6 to 15.6) | +0.01 (-0.10 to 0.12) |
| RoB-BERT minus Gemini 4 Argon | n/a | n/a | -0.7 (-3.5 to 2.2) | -0.02 (-0.08 to 0.04) | -5.7 (-13.5 to 1.6) | -0.07 (-0.19 to 0.04) |
| **Cochrane (42 rows)** | | | | | | |
| RoB-BERT minus Claude Fable 5.1 | n/a | n/a | +2.9 (-2.4 to 8.6) | +0.07 (-0.10 to 0.24) | -4.8 (-23.3 to 14.3) | -0.02 (-0.28 to 0.26) |
| RoB-BERT minus GPT-6 Astra | n/a | n/a | +23.3 (14.6 to 31.2) | +0.21 (0.08 to 0.33) | +35.7 (19.0 to 53.7) | +0.32 (0.12 to 0.54) |
| RoB-BERT minus Gemini 4 Argon | n/a | n/a | +3.8 (-2.4 to 11.4) | +0.11 (-0.04 to 0.25) | -2.4 (-14.6 to 9.8) | +0.01 (-0.17 to 0.20) |
| **Pooled unassisted cohorts (419 rows)** | | | | | | |
| RoB-BERT minus Claude Fable 5.1 | +2.9 (1.0 to 4.8) | +0.03 (0.01 to 0.06) | +3.0 (1.0 to 4.9) | +0.05 (0.01 to 0.08) | -1.2 (-7.2 to 4.5) | -0.01 (-0.10 to 0.08) |
| RoB-BERT minus GPT-6 Astra | +1.1 (-1.3 to 3.5) | +0.02 (-0.01 to 0.05) | +15.2 (13.1 to 17.3) | +0.08 (0.05 to 0.11) | +15.0 (9.5 to 20.8) | +0.13 (0.06 to 0.21) |
| RoB-BERT minus Gemini 4 Argon | +10.8 (7.9 to 13.8) | +0.08 (0.05 to 0.11) | +1.2 (-0.8 to 3.2) | +0.03 (-0.01 to 0.06) | -0.5 (-5.7 to 4.8) | +0.02 (-0.06 to 0.10) |
| **ROBoto2 assisted (151 rows)** | | | | | | |
| RoB-BERT minus Claude Fable 5.1 | +1.3 (-0.5 to 3.1) | +0.03 (0.00 to 0.05) | +1.3 (-1.5 to 4.2) | +0.03 (-0.03 to 0.08) | +5.3 (-4.0 to 15.2) | +0.11 (-0.05 to 0.26) |
| RoB-BERT minus GPT-6 Astra | +1.3 (-0.4 to 3.1) | +0.01 (-0.01 to 0.03) | +10.6 (7.4 to 13.5) | +0.07 (0.02 to 0.12) | +14.6 (4.6 to 24.5) | +0.23 (0.08 to 0.37) |

| Comparison | Signalling question Δ agreement, points (95% CI) | Signalling question Δκ (95% CI) | Domain Δ agreement, points (95% CI) | Domain Δκ (95% CI) | Overall Δ agreement, points (95% CI) | Overall Δκ (95% CI) |
|---|---|---|---|---|---|---|
| RoB-BERT minus Gemini 4 Argon | -2.3 (-4.9 to 0.3) | -0.01 (-0.04 to 0.02) | -0.1 (-2.9 to 2.8) | -0.01 (-0.05 to 0.04) | +4.0 (-4.6 to 12.6) | +0.05 (-0.09 to 0.19) |

Positive values favour RoB-BERT. Each pair is scored on identical trial-level bootstrap resamples (2,000 replicates, seed 20261103).

### Supplementary Table S6. Direction of error by cohort, primary population.

| AI system | Overall judgement below the reference category, n (%; 95% CI) | Sensitivity for high-risk domains, % (95% CI) | Specificity for high-risk domains, % (95% CI) | Domains called high that the reference did not, n |
|---|---|---|---|---|
| **ROBoto2 unassisted (185 rows; 50 high-risk reference domains)** | | | | |
| RoB-BERT v6.1 | 21 (11.4; 7.0 to 16.2) | 24.0 (13.5 to 34.0) | 90.9 (88.9 to 92.8) | 80 |
| RoB-BERT reader alone | 21 (11.4; 7.0 to 16.2) | 26.0 (15.2 to 36.7) | 90.6 (88.6 to 92.7) | 82 |
| RoB-BERT encoder alone | 31 (16.8; 11.4 to 22.7) | 12.0 (4.3 to 20.0) | 93.8 (92.1 to 95.5) | 54 |
| Claude Fable 5.1 | 42 (22.7; 16.8 to 28.6) | 24.0 (11.8 to 37.0) | 91.0 (88.5 to 93.3) | 79 |
| GPT-6 Astra | 11 (5.9; 2.7 to 9.7) | 34.0 (18.6 to 48.1) | 79.7 (76.8 to 82.4) | 178 |
| Gemini 4 Argon | 47 (25.4; 19.5 to 31.9) | 22.0 (10.0 to 34.4) | 93.5 (91.4 to 95.4) | 57 |
| **LogiMed-RoB (192 rows; 61 high-risk reference domains)** | | | | |
| RoB-BERT v6.1 | 46 (24.0; 18.2 to 30.2) | 18.0 (9.4 to 27.9) | 93.7 (91.8 to 95.4) | 57 |
| RoB-BERT reader alone | 45 (23.4; 17.7 to 29.7) | 18.0 (9.4 to 27.9) | 93.2 (91.3 to 95.0) | 61 |
| RoB-BERT encoder alone | 45 (23.4; 17.7 to 29.7) | 3.3 (0.0 to 8.1) | 96.4 (95.1 to 97.6) | 32 |
| Claude Fable 5.1 | 46 (24.0; 18.2 to 30.2) | 21.3 (10.7 to 33.3) | 93.4 (91.3 to 95.5) | 59 |
| GPT-6 Astra | 9 (4.7; 2.1 to 7.8) | 45.9 (34.9 to 58.0) | 79.9 (77.1 to 82.6) | 181 |
| Gemini 4 Argon | 47 (24.5; 18.7 to 30.7) | 24.6 (12.5 to 37.7) | 95.7 (94.0 to 97.2) | 39 |
| **Cochrane (42 rows; 6 high-risk reference domains)** | | | | |
| RoB-BERT v6.1 | 6 (14.3; 4.7 to 25.6) | 50.0 (0.0 to 100.0) | 94.6 (91.0 to 97.6) | 11 |
| RoB-BERT reader alone | 6 (14.3; 4.7 to 25.6) | 50.0 (0.0 to 100.0) | 94.6 (91.0 to 97.6) | 11 |
| RoB-BERT encoder alone | 11 (26.2; 14.0 to 39.0) | 0.0 (0.0 to 0.0) | 99.5 (98.5 to 100.0) | 1 |
| Claude Fable 5.1 | 6 (14.3; 4.8 to 25.6) | 33.3 (0.0 to 80.0) | 97.5 (94.8 to 99.5) | 5 |
| GPT-6 Astra | 1 (2.4; 0.0 to 7.3) | 66.7 (0.0 to 100.0) | 81.9 (77.4 to 86.2) | 37 |
| Gemini 4 Argon | 9 (21.4; 9.5 to 34.1) | 50.0 (0.0 to 100.0) | 98.5 (96.0 to 100.0) | 3 |
| **Pooled unassisted cohorts (419 rows; 117 high-risk reference domains)** | | | | |
| RoB-BERT v6.1 | 73 (17.4; 13.8 to 20.8) | 22.2 (15.7 to 29.2) | 92.5 (91.2 to 93.7) | 148 |

| AI system | Overall judgement below the reference category, n (%; 95% CI) | Sensitivity for high-risk domains, % (95% CI) | Specificity for high-risk domains, % (95% CI) | Domains called high that the reference did not, n |
|---|---|---|---|---|
| RoB-BERT reader alone | 72 (17.2; 13.6 to 20.8) | 23.1 (16.4 to 30.0) | 92.2 (90.9 to 93.5) | 154 |
| RoB-BERT encoder alone | 87 (20.8; 16.9 to 24.7) | 6.8 (2.7 to 11.0) | 95.6 (94.7 to 96.5) | 87 |
| Claude Fable 5.1 | 94 (22.4; 18.5 to 26.3) | 23.1 (14.9 to 32.0) | 92.8 (91.3 to 94.3) | 143 |
| GPT-6 Astra | 21 (5.0; 3.1 to 7.2) | 41.9 (33.0 to 51.3) | 80.0 (78.3 to 81.8) | 396 |
| Gemini 4 Argon | 103 (24.6; 20.3 to 28.5) | 24.8 (16.4 to 33.3) | 95.0 (93.8 to 96.1) | 99 |
| **ROBoto2 assisted (151 rows; 80 high-risk reference domains)** | | | | |
| RoB-BERT v6.1 | 31 (20.5; 13.9 to 27.8) | 13.8 (5.4 to 23.5) | 92.3 (90.4 to 94.0) | 52 |
| RoB-BERT reader alone | 32 (21.2; 14.6 to 27.8) | 15.0 (6.2 to 24.6) | 92.1 (90.2 to 93.9) | 53 |
| RoB-BERT encoder alone | 46 (30.5; 23.2 to 37.7) | 5.0 (1.1 to 10.2) | 97.6 (96.4 to 98.7) | 16 |
| Claude Fable 5.1 | 45 (29.8; 22.5 to 37.7) | 13.8 (4.5 to 25.0) | 93.3 (91.0 to 95.5) | 45 |
| GPT-6 Astra | 17 (11.3; 6.6 to 16.6) | 26.2 (15.8 to 36.2) | 81.3 (78.6 to 84.1) | 126 |
| Gemini 4 Argon | 48 (31.8; 23.8 to 39.7) | 15.0 (6.2 to 24.7) | 95.1 (93.1 to 96.9) | 33 |

An invalid output has no judgement and counts as not high. Cochrane intervals for sensitivity are wide because the reference contains six high-risk domains.

### Supplementary Table S7. Component analyses: combined system minus reader alone and minus encoder alone.

| Comparison | Signalling question Δ agreement, points (95% CI) | Signalling question Δκ (95% CI) | Domain Δ agreement, points (95% CI) | Domain Δκ (95% CI) | Overall Δ agreement, points (95% CI) | Overall Δκ (95% CI) |
|---|---|---|---|---|---|---|
| **ROBoto2 unassisted (185 rows)** | | | | | | |
| RoB-BERT minus reader alone | +1.0 (0.0 to 1.9) | +0.01 (-0.00 to 0.02) | -0.2 (-1.0 to 0.6) | -0.00 (-0.02 to 0.01) | +0.0 (-2.2 to 2.2) | -0.01 (-0.04 to 0.02) |
| RoB-BERT minus encoder alone | -0.8 (-2.4 to 0.9) | +0.00 (-0.02 to 0.02) | -0.9 (-3.1 to 1.5) | -0.01 (-0.05 to 0.03) | +6.5 (-2.2 to 14.6) | +0.24 (0.10 to 0.37) |
| **Pooled unassisted cohorts (419 rows)** | | | | | | |
| RoB-BERT minus reader alone | +1.0 (0.1 to 1.9) | +0.01 (-0.00 to 0.02) | -0.3 (-0.8 to 0.2) | -0.00 (-0.01 to 0.01) | -0.5 (-1.7 to 0.5) | -0.01 (-0.03 to 0.00) |
| RoB-BERT minus encoder alone | -0.8 (-2.5 to 0.9) | +0.00 (-0.02 to 0.02) | +0.9 (-0.9 to 2.6) | +0.02 (-0.01 to 0.05) | +1.7 (-4.3 to 7.4) | +0.11 (0.02 to 0.20) |

The combined system uses the reader's answers with the encoder's answer substituted wherever the encoder's probability is at least 0.85; the gate replaced 550 of 14,752 reader answers on the benchmark.

**Supplementary Table S8. Sensitivity analysis SA1: rows with a valid output from every AI system (559 of 570 primary rows).**

| AI system | Valid/rows | Signalling question agreement, % (95% CI) | Signalling question κ (95% CI) | Domain agreement, % (95% CI) | Domain κ (95% CI) | Overall agreement, % (95% CI) | Overall κ (95% CI) |
|---|---|---|---|---|---|---|---|
| **ROBoto2 unassisted (184 rows, 184 trials)** | | | | | | | |
| RoB-BERT v6.1 | 184/184 | 41.8 (39.9 to 44.0) | 0.26 (0.24 to 0.29) | 67.2 (63.9 to 70.4) | 0.35 (0.29 to 0.40) | 53.3 (46.2 to 60.3) | 0.20 (0.09 to 0.31) |
| RoB-BERT reader alone | 184/184 | 40.9 (38.7 to 43.2) | 0.25 (0.23 to 0.28) | 67.4 (64.1 to 70.8) | 0.35 (0.30 to 0.41) | 53.3 (46.2 to 60.3) | 0.21 (0.10 to 0.31) |
| RoB-BERT encoder alone | 184/184 | 42.6 (40.9 to 44.5) | 0.26 (0.24 to 0.28) | 68.2 (65.2 to 71.2) | 0.36 (0.31 to 0.41) | 47.3 (40.2 to 54.3) | -0.03 (-0.14 to 0.08) |
| Claude Fable 5.1 | 184/184 | 38.9 (36.9 to 40.9) | 0.23 (0.20 to 0.25) | 62.3 (58.7 to 65.7) | 0.25 (0.20 to 0.31) | 46.2 (39.1 to 53.3) | 0.09 (-0.02 to 0.21) |
| GPT-6 Astra | 184/184 | 40.6 (38.5 to 42.8) | 0.24 (0.22 to 0.27) | 55.5 (52.1 to 58.9) | 0.27 (0.22 to 0.31) | 34.8 (28.3 to 41.8) | 0.02 (-0.06 to 0.09) |
| Gemini 4 Argon | 184/184 | 31.1 (28.3 to 34.2) | 0.18 (0.16 to 0.21) | 65.0 (61.7 to 68.2) | 0.29 (0.24 to 0.35) | 48.4 (41.3 to 56.0) | 0.09 (-0.03 to 0.21) |
| **Pooled unassisted cohorts (410 rows, 409 trials)** | | | | | | | |
| RoB-BERT v6.1 | 410/410 | 41.8 (39.8 to 43.9) | 0.26 (0.24 to 0.29) | 67.8 (65.6 to 70.1) | 0.29 (0.25 to 0.33) | 46.6 (41.6 to 51.5) | 0.17 (0.09 to 0.24) |
| RoB-BERT reader alone | 410/410 | 40.9 (38.6 to 43.1) | 0.25 (0.23 to 0.28) | 68.0 (65.8 to 70.4) | 0.29 (0.25 to 0.33) | 47.1 (42.0 to 52.1) | 0.18 (0.10 to 0.25) |
| RoB-BERT encoder alone | 410/410 | 42.6 (40.8 to 44.5) | 0.26 (0.24 to 0.29) | 66.9 (64.9 to 69.1) | 0.27 (0.23 to 0.31) | 44.9 (40.0 to 49.8) | 0.06 (-0.02 to 0.13) |
| Claude Fable 5.1 | 410/410 | 38.9 (36.7 to 41.0) | 0.23 (0.20 to 0.25) | 65.7 (63.3 to 68.1) | 0.25 (0.20 to 0.29) | 48.0 (43.2 to 53.2) | 0.18 (0.10 to 0.25) |
| GPT-6 Astra | 410/410 | 40.6 (38.3 to 42.8) | 0.24 (0.21 to 0.27) | 53.1 (51.0 to 55.3) | 0.21 (0.18 to 0.24) | 31.0 (26.6 to 35.5) | 0.03 (-0.02 to 0.07) |
| Gemini 4 Argon | 410/410 | 31.1 (28.2 to 34.2) | 0.18 (0.16 to 0.21) | 67.7 (65.5 to 70.0) | 0.27 (0.23 to 0.31) | 47.6 (42.5 to 52.3) | 0.15 (0.07 to 0.23) |
| **Paired differences, pooled unassisted cohorts** | | | | | | | |
| RoB-BERT minus Claude Fable 5.1 | | +3.0 (1.1 to 4.9) | +0.03 (0.01 to 0.06) | +2.1 (0.1 to 4.0) | +0.04 (0.00 to 0.08) | -1.5 (-7.3 to 4.4) | -0.01 (-0.09 to 0.08) |
| RoB-BERT minus GPT-6 Astra | | +1.2 (-1.1 to 3.5) | +0.02 (-0.01 to 0.05) | +14.7 (12.6 to 16.9) | +0.08 (0.04 to 0.11) | +15.6 (10.0 to 21.1) | +0.14 (0.06 to 0.22) |
| RoB-BERT minus Gemini 4 Argon | | +10.7 (7.8 to 13.6) | +0.08 (0.05 to 0.11) | +0.1 (-1.7 to 1.9) | +0.02 (-0.02 to 0.06) | -1.0 (-6.1 to 4.4) | +0.02 (-0.07 to 0.10) |

In the paired-difference rows the columns give the difference in agreement (points) and in κ with 95% intervals.

**Supplementary Table S9. Sensitivity analysis SA2: all outcome rows with text (1,008 rows; 857 pooled unassisted rows).**

| AI system | Valid/rows | Domain agreement, % (95% CI) | Domain κ (95% CI) | Overall agreement, % (95% CI) | Overall κ (95% CI) |
|---|---|---|---|---|---|
| **LogiMed-RoB (600 rows, 192 trials)** | | | | | |
| RoB-BERT v6.1 | 599/600 | 67.1 (62.7 to 71.1) | 0.16 (0.10 to 0.23) | 38.5 (31.5 to 45.7) | 0.07 (-0.03 to 0.18) |
| RoB-BERT reader alone | 599/600 | 67.0 (62.7 to 71.2) | 0.16 (0.10 to 0.23) | 39.5 (32.6 to 46.6) | 0.09 (-0.01 to 0.19) |
| RoB-BERT encoder alone | 600/600 | 64.9 (60.2 to 69.5) | 0.12 (0.05 to 0.20) | 41.0 (32.7 to 49.5) | 0.07 (-0.05 to 0.19) |
| Claude Fable 5.1 | 593/600 | 66.8 (62.2 to 71.4) | 0.18 (0.11 to 0.25) | 45.3 (38.6 to 51.8) | 0.16 (0.06 to 0.25) |
| GPT-6 Astra | 600/600 | 49.6 (46.4 to 52.9) | 0.13 (0.08 to 0.18) | 27.5 (20.9 to 33.9) | 0.01 (-0.05 to 0.07) |
| Gemini 4 Argon | 594/600 | 69.4 (65.6 to 73.2) | 0.21 (0.14 to 0.28) | 43.8 (36.8 to 50.7) | 0.13 (0.02 to 0.24) |
| **Cochrane (72 rows, 41 trials)** | | | | | |
| RoB-BERT v6.1 | 72/72 | 85.3 (81.2 to 89.3) | 0.45 (0.31 to 0.57) | 58.3 (44.3 to 72.1) | 0.32 (0.10 to 0.53) |
| RoB-BERT reader alone | 72/72 | 84.4 (80.3 to 88.4) | 0.40 (0.28 to 0.51) | 56.9 (43.4 to 70.4) | 0.30 (0.10 to 0.50) |
| RoB-BERT encoder alone | 72/72 | 84.2 (79.4 to 88.6) | 0.37 (0.24 to 0.47) | 61.1 (46.5 to 74.7) | 0.32 (0.11 to 0.52) |
| Claude Fable 5.1 | 72/72 | 81.4 (75.9 to 86.3) | 0.35 (0.22 to 0.46) | 55.6 (40.8 to 69.2) | 0.25 (0.04 to 0.45) |
| GPT-6 Astra | 72/72 | 59.2 (52.9 to 65.2) | 0.21 (0.12 to 0.31) | 20.8 (10.7 to 31.9) | -0.04 (-0.13 to 0.07) |
| Gemini 4 Argon | 69/72 | 81.9 (76.3 to 87.1) | 0.33 (0.18 to 0.47) | 58.3 (45.1 to 71.4) | 0.30 (0.07 to 0.52) |
| **Pooled unassisted cohorts (857 rows, 418 trials)** | | | | | |
| RoB-BERT v6.1 | 856/857 | 68.7 (65.5 to 71.7) | 0.24 (0.19 to 0.29) | 43.4 (38.0 to 48.7) | 0.14 (0.06 to 0.22) |
| RoB-BERT reader alone | 856/857 | 68.6 (65.4 to 71.6) | 0.24 (0.19 to 0.28) | 44.0 (38.7 to 49.2) | 0.15 (0.07 to 0.23) |
| RoB-BERT encoder alone | 857/857 | 67.2 (63.9 to 70.4) | 0.21 (0.15 to 0.26) | 44.0 (38.0 to 50.0) | 0.09 (-0.00 to 0.18) |
| Claude Fable 5.1 | 850/857 | 67.0 (63.4 to 70.5) | 0.22 (0.17 to 0.27) | 46.4 (41.3 to 51.5) | 0.17 (0.09 to 0.24) |
| GPT-6 Astra | 857/857 | 51.7 (49.1 to 54.2) | 0.17 (0.13 to 0.20) | 28.6 (23.6 to 33.5) | 0.01 (-0.04 to 0.06) |
| Gemini 4 Argon | 847/857 | 69.4 (66.4 to 72.2) | 0.25 (0.20 to 0.29) | 46.0 (40.9 to 51.1) | 0.15 (0.07 to 0.23) |
| **Paired differences, pooled unassisted cohorts (857 rows)** | | | | | |
| RoB-BERT minus Claude Fable 5.1 | | +1.6 (-0.4 to 3.8) | +0.02 (-0.02 to 0.06) | -3.0 (-8.9 to 2.9) | -0.03 (-0.11 to 0.06) |
| RoB-BERT minus GPT-6 Astra | | +16.9 (14.0 to 19.8) | +0.07 (0.03 to 0.11) | +14.8 (8.4 to 21.1) | +0.13 (0.05 to 0.22) |
| RoB-BERT minus Gemini 4 Argon | | -0.8 (-2.8 to 1.1) | -0.01 (-0.05 to 0.03) | -2.6 (-8.3 to 2.7) | -0.01 (-0.09 to 0.07) |

ROBoto2 rows are study-level and are unchanged from the primary analysis. Signalling-question results are identical to the primary analysis and are omitted.

**Supplementary Table S10. Sensitivity analysis SA3: articles truncated by the reader's preprocessing versus not truncated, primary population.**

| AI system | Valid/rows | Domain agreement, % (95% CI) | Domain κ (95% CI) | Overall agreement, % (95% CI) | Overall κ (95% CI) |
|---|---|---|---|---|---|
| **Truncated by the reader's preprocessing: LogiMed-RoB (77 rows, 77 trials)** | | | | | |
| RoB-BERT v6.1 | 77/77 | 64.7 (59.2 to 70.1) | -0.02 (-0.09 to 0.06) | 24.7 (15.6 to 33.8) | -0.15 (-0.29 to -0.02) |
| RoB-BERT reader alone | 77/77 | 65.5 (60.0 to 70.9) | -0.01 (-0.09 to 0.07) | 26.0 (16.9 to 35.1) | -0.13 (-0.27 to 0.01) |
| RoB-BERT encoder alone | 77/77 | 61.6 (55.8 to 67.3) | -0.01 (-0.10 to 0.08) | 32.5 (22.1 to 42.9) | -0.08 (-0.22 to 0.08) |
| Claude Fable 5.1 | 76/77 | 65.5 (59.2 to 71.4) | 0.01 (-0.07 to 0.10) | 41.6 (31.2 to 53.2) | 0.07 (-0.10 to 0.23) |
| GPT-6 Astra | 77/77 | 49.4 (44.9 to 54.0) | 0.07 (0.01 to 0.14) | 26.0 (16.9 to 36.4) | -0.01 (-0.11 to 0.10) |
| Gemini 4 Argon | 75/77 | 65.5 (59.7 to 71.2) | 0.01 (-0.05 to 0.09) | 35.1 (24.7 to 45.5) | -0.01 (-0.17 to 0.15) |
| **Truncated by the reader's preprocessing: Cochrane (8 rows, 8 trials)** | | | | | |
| RoB-BERT v6.1 | 8/8 | 85.0 (80.0 to 92.5) | 0.35 (-0.04 to 0.64) | 25.0 (0.0 to 62.5) | 0.00 (-0.23 to 0.27) |
| RoB-BERT reader alone | 8/8 | 85.0 (80.0 to 92.5) | 0.35 (-0.04 to 0.64) | 25.0 (0.0 to 62.5) | 0.00 (-0.23 to 0.27) |
| RoB-BERT encoder alone | 8/8 | 85.0 (72.5 to 95.0) | 0.19 (-0.08 to 0.53) (8 undefined) | 62.5 (25.0 to 87.5) | 0.27 (-0.26 to 0.76) (8 undefined) |
| Claude Fable 5.1 | 8/8 | 90.0 (82.5 to 97.5) | 0.52 (0.15 to 0.79) (1 undefined) | 62.5 (25.0 to 100.0) | 0.38 (0.00 to 1.00) (1 undefined) |
| GPT-6 Astra | 8/8 | 57.5 (50.0 to 65.0) | 0.14 (0.01 to 0.30) | 12.5 (0.0 to 37.5) | 0.00 (0.00 to 0.00) |
| Gemini 4 Argon | 8/8 | 92.5 (85.0 to 97.5) | 0.54 (-0.03 to 0.88) (4 undefined) | 75.0 (37.5 to 100.0) | 0.53 (-0.23 to 1.00) (4 undefined) |
| **Truncated by the reader's preprocessing: Pooled unassisted cohorts (87 rows, 87 trials)** | | | | | |
| RoB-BERT v6.1 | 87/87 | 66.0 (60.9 to 71.0) | 0.04 (-0.04 to 0.14) | 24.1 (14.9 to 33.3) | -0.14 (-0.27 to -0.01) |
| RoB-BERT reader alone | 87/87 | 66.9 (61.8 to 72.0) | 0.05 (-0.04 to 0.15) | 25.3 (16.1 to 34.5) | -0.12 (-0.25 to 0.01) |
| RoB-BERT encoder alone | 87/87 | 63.4 (58.2 to 68.7) | 0.03 (-0.06 to 0.11) | 34.5 (25.3 to 44.8) | -0.04 (-0.18 to 0.10) |
| Claude Fable 5.1 | 86/87 | 67.4 (61.8 to 72.9) | 0.06 (-0.02 to 0.14) | 43.7 (33.3 to 54.0) | 0.09 (-0.06 to 0.26) |
| GPT-6 Astra | 87/87 | 50.3 (46.2 to 54.5) | 0.09 (0.03 to 0.15) | 24.1 (14.9 to 33.3) | -0.01 (-0.10 to 0.09) |
| Gemini 4 Argon | 85/87 | 67.6 (62.1 to 72.9) | 0.06 (-0.02 to 0.14) | 39.1 (29.9 to 49.4) | 0.05 (-0.11 to 0.21) |
| **Truncated by the reader's preprocessing: paired differences, pooled unassisted cohorts** | | | | | |
| RoB-BERT minus Claude Fable 5.1 | | -1.4 (-4.6 to 2.3) | -0.01 (-0.09 to 0.07) | -19.5 (-31.0 to -8.0) | -0.23 (-0.41 to -0.05) |
| RoB-BERT minus GPT-6 Astra | | +15.6 (11.0 to 20.2) | -0.04 (-0.13 to 0.04) | +0.0 (-12.6 to 12.6) | -0.13 (-0.28 to 0.03) |
| RoB-BERT minus Gemini 4 Argon | | -1.6 (-5.5 to 2.3) | -0.01 (-0.08 to 0.05) | -14.9 (-26.4 to -4.6) | -0.19 (-0.35 to -0.02) |
| **Not truncated: LogiMed-RoB (115 rows, 115 trials)** | | | | | |

| AI system | Valid/rows | Domain agreement, % (95% CI) | Domain κ (95% CI) | Overall agreement, % (95% CI) | Overall κ (95% CI) |
|---|---|---|---|---|---|
| RoB-BERT v6.1 | 115/115 | 65.2 (60.7 to 69.6) | 0.24 (0.17 to 0.32) | 44.3 (34.8 to 53.9) | 0.16 (0.03 to 0.30) |
| RoB-BERT reader alone | 115/115 | 65.7 (61.6 to 70.1) | 0.25 (0.17 to 0.32) | 45.2 (35.7 to 54.8) | 0.18 (0.05 to 0.31) |
| RoB-BERT encoder alone | 115/115 | 63.0 (59.3 to 66.8) | 0.19 (0.12 to 0.26) | 41.7 (33.0 to 51.3) | 0.10 (0.00 to 0.21) |
| Claude Fable 5.1 | 113/115 | 63.3 (57.9 to 68.5) | 0.25 (0.17 to 0.33) | 47.0 (38.3 to 56.5) | 0.21 (0.08 to 0.34) |
| GPT-6 Astra | 115/115 | 47.5 (43.3 to 51.5) | 0.15 (0.10 to 0.21) | 31.3 (23.5 to 40.0) | 0.04 (-0.04 to 0.12) |
| Gemini 4 Argon | 114/115 | 65.9 (61.4 to 70.4) | 0.27 (0.19 to 0.34) | 47.0 (37.4 to 56.5) | 0.19 (0.06 to 0.32) |
| **Not truncated: Cochrane (34 rows, 33 trials)** | | | | | |
| RoB-BERT v6.1 | 34/34 | 83.5 (77.6 to 89.1) | 0.51 (0.37 to 0.64) | 64.7 (47.1 to 81.8) | 0.43 (0.15 to 0.70) |
| RoB-BERT reader alone | 34/34 | 82.4 (75.8 to 88.5) | 0.46 (0.32 to 0.60) | 64.7 (47.1 to 81.8) | 0.43 (0.15 to 0.70) |
| RoB-BERT encoder alone | 34/34 | 82.4 (77.0 to 87.3) | 0.44 (0.31 to 0.53) | 61.8 (44.1 to 77.1) | 0.33 (0.06 to 0.57) |
| Claude Fable 5.1 | 34/34 | 78.8 (71.8 to 85.5) | 0.40 (0.24 to 0.54) | 61.8 (44.4 to 78.8) | 0.36 (0.08 to 0.63) |
| GPT-6 Astra | 34/34 | 61.2 (53.7 to 69.1) | 0.30 (0.20 to 0.42) | 23.5 (11.1 to 39.4) | 0.01 (-0.12 to 0.16) |
| Gemini 4 Argon | 32/34 | 77.1 (67.6 to 85.3) | 0.36 (0.18 to 0.54) | 55.9 (38.2 to 72.7) | 0.29 (-0.00 to 0.55) |
| **Not truncated: Pooled unassisted cohorts (332 rows, 331 trials)** | | | | | |
| RoB-BERT v6.1 | 332/332 | 68.4 (66.0 to 70.8) | 0.33 (0.29 to 0.37) | 51.8 (46.2 to 57.1) | 0.23 (0.14 to 0.31) |
| RoB-BERT reader alone | 332/332 | 68.6 (66.0 to 70.9) | 0.33 (0.29 to 0.37) | 52.1 (46.5 to 57.5) | 0.23 (0.15 to 0.32) |
| RoB-BERT encoder alone | 332/332 | 68.0 (65.7 to 70.0) | 0.31 (0.27 to 0.35) | 47.0 (41.4 to 52.4) | 0.06 (-0.02 to 0.15) |
| Claude Fable 5.1 | 330/332 | 64.3 (61.5 to 67.0) | 0.26 (0.22 to 0.31) | 48.2 (42.8 to 53.5) | 0.17 (0.09 to 0.25) |
| GPT-6 Astra | 332/332 | 53.4 (51.0 to 56.0) | 0.23 (0.20 to 0.27) | 32.8 (27.9 to 37.8) | 0.04 (-0.02 to 0.09) |
| Gemini 4 Argon | 328/332 | 66.4 (64.0 to 68.9) | 0.29 (0.25 to 0.33) | 48.5 (42.9 to 53.8) | 0.15 (0.06 to 0.24) |
| **Not truncated: paired differences, pooled unassisted cohorts** | | | | | |
| RoB-BERT minus Claude Fable 5.1 | | +4.1 (1.9 to 6.4) | +0.07 (0.02 to 0.11) | +3.6 (-3.3 to 9.6) | +0.05 (-0.05 to 0.15) |
| RoB-BERT minus GPT-6 Astra | | +15.1 (12.4 to 17.6) | +0.10 (0.06 to 0.14) | +19.0 (12.7 to 25.1) | +0.19 (0.10 to 0.28) |
| RoB-BERT minus Gemini 4 Argon | | +2.0 (-0.3 to 4.3) | +0.04 (-0.01 to 0.08) | +3.3 (-2.4 to 9.3) | +0.07 (-0.02 to 0.17) |

Two ROBoto2 unassisted rows were truncated and are included in the pooled rows but not tabulated separately. The frontier models received the full article in every case. Intervals marked undefined exclude bootstrap replicates in which κ could not be computed.

**Supplementary Table S11. Sensitivity analysis SA4: Cochrane cohort without the RECOVERY platform trial comparison (NCT04381936).**

| AI system | Valid/rows | Domain agreement, % (95% CI) | Domain κ (95% CI) | Overall agreement, % (95% CI) | Overall κ (95% CI) |
|---|---|---|---|---|---|
| **Cochrane (41 rows, 40 trials)** | | | | | |
| RoB-BERT v6.1 | 41/41 | 83.9 (79.0 to 88.4) | 0.50 (0.36 to 0.61) | 58.5 (42.9 to 72.5) | 0.36 (0.13 to 0.58) |
| RoB-BERT reader alone | 41/41 | 82.9 (77.5 to 87.7) | 0.46 (0.32 to 0.58) | 58.5 (42.9 to 72.5) | 0.36 (0.13 to 0.58) |
| RoB-BERT encoder alone | 41/41 | 82.9 (78.0 to 87.5) | 0.41 (0.29 to 0.52) | 63.4 (47.6 to 78.0) | 0.34 (0.08 to 0.58) |
| Claude Fable 5.1 | 41/41 | 81.0 (74.8 to 86.7) | 0.42 (0.29 to 0.55) | 63.4 (48.8 to 78.0) | 0.38 (0.13 to 0.61) |
| GPT-6 Astra | 41/41 | 60.0 (53.5 to 66.3) | 0.28 (0.18 to 0.38) | 22.0 (9.8 to 35.0) | 0.05 (-0.04 to 0.15) |
| Gemini 4 Argon | 39/41 | 80.0 (71.6 to 86.8) | 0.39 (0.21 to 0.55) | 61.0 (45.2 to 75.6) | 0.35 (0.07 to 0.60) |
| **Cochrane: paired differences** | | | | | |
| RoB-BERT minus Claude Fable 5.1 | | +2.9 (-2.5 to 8.5) | +0.08 (-0.09 to 0.23) | -4.9 (-24.4 to 12.5) | -0.01 (-0.29 to 0.25) |
| RoB-BERT minus GPT-6 Astra | | +23.9 (15.6 to 31.9) | +0.22 (0.09 to 0.33) | +36.6 (19.5 to 53.7) | +0.32 (0.09 to 0.54) |
| RoB-BERT minus Gemini 4 Argon | | +3.9 (-2.4 to 11.5) | +0.11 (-0.04 to 0.25) | -2.4 (-15.0 to 9.8) | +0.01 (-0.16 to 0.20) |
| **Pooled unassisted cohorts (418 rows, 417 trials)** | | | | | |
| RoB-BERT v6.1 | 418/418 | 67.9 (65.5 to 70.2) | 0.29 (0.25 to 0.33) | 46.2 (41.2 to 51.1) | 0.16 (0.09 to 0.23) |
| RoB-BERT reader alone | 418/418 | 68.2 (65.8 to 70.5) | 0.29 (0.25 to 0.33) | 46.7 (41.9 to 51.6) | 0.17 (0.10 to 0.24) |
| RoB-BERT encoder alone | 418/418 | 67.0 (64.7 to 69.1) | 0.27 (0.23 to 0.31) | 44.5 (39.7 to 49.2) | 0.05 (-0.02 to 0.13) |
| Claude Fable 5.1 | 415/418 | 64.9 (62.5 to 67.4) | 0.24 (0.20 to 0.28) | 47.4 (42.6 to 51.8) | 0.17 (0.09 to 0.24) |
| GPT-6 Astra | 418/418 | 52.7 (50.6 to 54.7) | 0.21 (0.18 to 0.23) | 31.1 (26.9 to 35.6) | 0.03 (-0.01 to 0.08) |
| Gemini 4 Argon | 412/418 | 66.7 (64.4 to 69.0) | 0.26 (0.22 to 0.30) | 46.7 (41.6 to 51.7) | 0.14 (0.06 to 0.22) |
| **Pooled unassisted cohorts: paired differences** | | | | | |
| RoB-BERT minus Claude Fable 5.1 | | +3.0 (1.0 to 5.1) | +0.05 (0.01 to 0.09) | -1.2 (-6.9 to 4.8) | -0.01 (-0.10 to 0.08) |
| RoB-BERT minus GPT-6 Astra | | +15.2 (13.0 to 17.4) | +0.08 (0.05 to 0.12) | +15.1 (9.5 to 20.7) | +0.13 (0.05 to 0.21) |
| RoB-BERT minus Gemini 4 Argon | | +1.2 (-0.7 to 3.3) | +0.03 (-0.01 to 0.07) | -0.5 (-5.8 to 5.0) | +0.02 (-0.07 to 0.10) |

One primary row (cochrane:001, the aspirin comparison) was excluded; the baricitinib comparison has no open-access text and was never among the scored rows.

#### Supplementary Table S12. Descriptive context: human inter-assessor agreement and the earlier GPT-4 evaluation.

| Comparison | Signalling question agreement, % | Domain agreement, % | Overall agreement, % |
|---|---|---|---|
| Human: unassisted versus assisted assessor, ROBoto2 (52 pairs on 52 trials) | 42.0 (κ 0.26) | 61.2 (κ 0.15) | 46.2 (κ 0.17) |
| Human: assisted versus assisted assessor, ROBoto2 (60 pairs on 10 trials) | 46.0 (κ 0.30) | 64.7 (κ 0.35) | 50.0 (κ 0.15) |
| GPT-4 (2023), optimized prompt, PDF plus registration and protocol, Cochrane (42 rows) | n/a | 52.9 (κ 0.03) | 42.9 (κ 0.00) |
| RoB-BERT v6.1, text only, Cochrane (42 rows) | n/a | 83.8 (κ 0.49) | 57.1 (κ 0.34) |

Descriptive only; no formal comparison was made. Human figures are from the benchmark's inter-assessor analysis of ROBoto2 trials assessed by more than one person; every pair includes at least one assessor who saw LLM suggestions. High-risk sensitivity and specificity for the same pairs are in Supplementary Table S16.

#### Supplementary Table S13. Adjudication of disagreements by domain (unweighted counts, 188 domains).

| Domain | Domains adjudicated | RoB-BERT correct | Reference correct | Both defensible | Both wrong | Indeterminate |
|---|---|---|---|---|---|---|
| Domain 1, randomization | 46 | 32 (70%) | 5 (11%) | 4 (9%) | 4 (9%) | 1 (2%) |
| Domain 2, deviations from intended interventions | 33 | 11 (33%) | 6 (18%) | 11 (33%) | 4 (12%) | 1 (3%) |
| Domain 3, missing outcome data | 35 | 13 (37%) | 9 (26%) | 2 (6%) | 7 (20%) | 4 (11%) |
| Domain 4, measurement of the outcome | 28 | 11 (39%) | 7 (25%) | 9 (32%) | 0 (0%) | 1 (4%) |
| Domain 5, selection of the reported result | 46 | 31 (67%) | 8 (17%) | 6 (13%) | 0 (0%) | 1 (2%) |

Counts are unweighted across the three sampled cohorts and pool the judgements of two reviewers (TP and DZ), who divided the sample between them and adjudicated independently; each domain was adjudicated by one reviewer. Of the 98 domains in which the reference was judged wrong and RoB-BERT correct, 55 were a low-risk rating for Domain 1 without allocation concealment information or for Domain 5 without an accessible analysis plan. Fourteen of 16 RoB-BERT errors in Domain 3 were high-risk calls built on miscounted denominators.

#### Supplementary Table S14. Types of error attributed to the reference and to RoB-BERT v6.1 in the adjudicated disagreements.

| Error type | Reference | RoB-BERT v6.1 |
|---|---|---|
| RoB 2 guidance misapplied | 44 (32 major) | 16 (9 major) |
| Reported information missed | 14 (10 major) | 11 (3 major) |
| Too lenient on a borderline call | 25 (2 major) | 7 (0 major) |
| Too strict on a borderline call | 13 (2 major) | 4 (0 major) |
| Factual error (contradicted by the report) | 3 (1 major) | 7 (3 major) |

| Error type | Reference | RoB-BERT v6.1 |
| --- | --- | --- |
| Relied on information outside the report | 13 (6 major) | 2 (0 major) |
| Assessed a different outcome or result | 1 (0 major) | 4 (2 major) |
| Direction when one side was judged correct: too lenient / too strict | 68 / 30 | 20 / 15 |

Errors are counted for every domain in which a side was judged wrong, including domains where both were wrong. Major: Low versus High, or a clear rule violation that would change a certainty rating. Reliance on information outside the report is not necessarily an error by the reviewer, who may have consulted a registry or protocol; it is shown separately for that reason.

### Supplementary Table S15. Agreement between the benchmark reference and an independent Cochrane review team that assessed the same trial.

| Domain | Second review agreed with reference |
| --- | --- |
| Domain 1 (randomization) | 19/28 (68%) |
| Domain 2 (deviations; RoB 1 blinding of participants and personnel) | 4/15 (27%) |
| Domain 3 (missing outcome data) | 14/27 (52%) |
| Domain 4 (measurement; RoB 1 blinding of outcome assessment) | 15/24 (62%) |
| Domain 5 (selective reporting) | 9/18 (50%) |
| Independent RoB 2 re-assessment (McCann 2021) | Disagreed on Domains 1 and 4 and overall |

23 of the 157 benchmark Cochrane trials were included and assessed by a second, independent Cochrane review team (identity verified by DOI, PMID or title; reviews sharing authors excluded). 22 of the second assessments used RoB 1, mapped approximately to RoB 2 domains; RoB 1 penalizes lack of blinding more than RoB 2 does for the effect of assignment, so Domain 2 and 4 disagreement partly reflects the tool. Several cross-review links in the CoreSR Cochrane database were false matches and were excluded.

### Supplementary Table S16. High-risk judgements: sensitivity and specificity between human assessors, and between each system and the same assessors, on ROBoto2 trials rated by more than one assessor.

**(a) Unassisted assessor versus LLM-assisted assessor: 52 pairs on 52 trials. The unassisted assessment is the benchmark reference for these trials.**

| Level | Compared with the unassisted (reference) assessor and with the assisted assessor | Sensitivity vs unassisted assessor | Specificity vs unassisted assessor | Sensitivity vs assisted assessor | Specificity vs assisted assessor | PSA with unassisted assessor | PSA with assisted assessor |
| --- | --- | --- | --- | --- | --- | --- | --- |
| Domain | Second human assessor | 23.1 (0.0 to 50.0) [3/13] | 90.3 (86.2 to 94.2) | 11.1 (0.0 to 23.8) [3/27] | 95.7 (92.4 to 98.3) | 15.0 (0.0 to 29.3) | Same as previous column |
| Domain | RoB-BERT v6.1 | 30.8 (7.7 to 61.5) [4/13] | 92.7 (89.5 to 95.8) | 18.5 (5.0 to 32.1) [5/27] | 92.7 (89.5 to 95.5) | 22.9 (5.9 to 41.0) | 20.4 (5.6 to 33.3) |
| Domain | RoB-BERT reader alone | 30.8 (7.7 to 61.5) [4/13] | 92.7 (89.5 to 95.8) | 18.5 (5.0 to 32.1) [5/27] | 92.7 (89.5 to 95.5) | 22.9 (5.9 to 41.0) | 20.4 (5.6 to 33.3) |

| Level | Compared with the unassisted (reference) assessor and with the assisted assessor | Sensitivity vs unassisted assessor | Specificity vs unassisted assessor | Sensitivity vs assisted assessor | Specificity vs assisted assessor | PSA with unassisted assessor | PSA with assisted assessor |
|---|---|---|---|---|---|---|---|
| Domain | RoB-BERT encoder alone | 7.7 (0.0 to 27.3) [1/13] | 97.2 (95.1 to 98.8) | 7.4 (0.0 to 17.7) [2/27] | 97.4 (95.3 to 99.2) | 9.5 (0.0 to 29.6) | 11.4 (0.0 to 26.1) |
| Domain | Claude Fable 5.1 | 23.1 (0.0 to 50.0) [3/13] | 91.1 (86.0 to 95.5) | 18.5 (3.4 to 35.5) [5/27] | 91.4 (86.4 to 95.6) | 15.8 (0.0 to 33.3) | 19.2 (3.8 to 35.6) |
| Domain | GPT-6 Astra | 38.5 (13.3 to 66.7) [5/13] | 82.6 (77.6 to 87.4) | 25.9 (5.6 to 45.2) [7/27] | 82.4 (77.5 to 86.9) | 16.4 (4.2 to 29.0) | 18.7 (3.6 to 31.4) |
| Domain | Gemini 4 Argon | 23.1 (0.0 to 43.8) [3/13] | 93.1 (88.9 to 96.8) | 18.5 (0.0 to 37.5) [5/27] | 93.6 (89.1 to 97.1) | 18.2 (0.0 to 34.5) | 21.3 (0.0 to 40.7) |
| Overall | Second human assessor | 50.0 (16.7 to 83.3) [5/10] | 64.3 (48.8 to 78.3) | 25.0 (6.2 to 45.0) [5/20] | 84.4 (70.3 to 96.6) | 33.3 (8.7 to 53.3) | Same as previous column |
| Overall | RoB-BERT v6.1 | 50.0 (16.7 to 83.3) [5/10] | 66.7 (51.3 to 81.0) | 50.0 (26.7 to 73.3) [10/20] | 71.9 (55.2 to 87.9) | 34.5 (10.0 to 57.1) | 51.3 (29.4 to 69.4) |
| Overall | RoB-BERT reader alone | 50.0 (16.7 to 83.3) [5/10] | 66.7 (51.3 to 81.0) | 50.0 (26.7 to 73.3) [10/20] | 71.9 (55.2 to 87.9) | 34.5 (10.0 to 57.1) | 51.3 (29.4 to 69.4) |
| Overall | RoB-BERT encoder alone | 10.0 (0.0 to 33.3) [1/10] | 83.3 (71.4 to 93.5) | 20.0 (4.5 to 38.9) [4/20] | 87.5 (75.0 to 97.1) | 11.1 (0.0 to 32.0) | 28.6 (7.4 to 50.0) |
| Overall | Claude Fable 5.1 | 40.0 (10.0 to 75.0) [4/10] | 71.4 (57.5 to 84.1) | 45.0 (23.8 to 66.7) [9/20] | 78.1 (62.5 to 91.2) | 30.8 (7.7 to 53.3) | 50.0 (27.6 to 68.0) |
| Overall | GPT-6 Astra | 90.0 (66.7 to 100.0) [9/10] | 42.9 (28.9 to 58.1) | 70.0 (50.0 to 89.5) [14/20] | 40.6 (24.1 to 58.1) | 41.9 (21.6 to 59.1) | 52.8 (34.0 to 68.8) |
| Overall | Gemini 4 Argon | 30.0 (0.0 to 62.5) [3/10] | 81.0 (68.9 to 91.7) | 35.0 (14.3 to 55.6) [7/20] | 87.5 (75.0 to 97.1) | 28.6 (0.0 to 52.6) | 45.2 (21.0 to 64.9) |

**(b) Two LLM-assisted assessors: 60 pairs on 10 trials (every assessor combination). Exploratory because of the small number of trials.**

| Level | Compared with assessor 1 and with assessor 2 | Sensitivity vs assessor 1 | Specificity vs assessor 1 | Sensitivity vs assessor 2 | Specificity vs assessor 2 | PSA with assessor 1 | PSA with assessor 2 |
|---|---|---|---|---|---|---|---|
| Domain | Second human assessor | 33.3 (12.5 to 47.8) [11/33] | 92.9 (89.1 to 96.1) | 36.7 (14.3 to 56.4) [11/30] | 91.9 (88.2 to 95.1) | 34.9 (13.3 to 51.3) | Same as previous column |
| Domain | RoB-BERT v6.1 | 42.4 (0.0 to 64.1) [14/33] | 94.0 (88.6 to 98.6) | 43.3 (0.0 to 66.7) [13/30] | 93.7 (88.0 to 98.2) | 44.4 (0.0 to 61.7) | 43.3 (0.0 to 59.2) |
| Domain | RoB-BERT reader alone | 42.4 (0.0 to 64.1) [14/33] | 91.8 (82.3 to 97.9) | 43.3 (0.0 to 66.7) [13/30] | 91.5 (81.7 to 97.9) | 40.6 (0.0 to 52.4) | 39.4 (0.0 to 50.0) |
| Domain | RoB-BERT encoder alone | 21.2 (0.0 to 42.4) [7/33] | 98.1 (94.3 to 100.0) | 26.7 (0.0 to 56.2) [8/30] | 98.5 (95.5 to 100.0) | 31.1 (0.0 to 56.2) | 38.1 (0.0 to 65.0) |
| Domain | Claude Fable 5.1 | 9.1 (0.0 to 31.6) [3/33] | 94.4 (88.7 to 100.0) | 10.0 (0.0 to 30.0) [3/30] | 94.4 (88.9 to 100.0) | 11.8 (0.0 to 32.1) | 12.5 (0.0 to 31.6) |

| Level | Compared with assessor 1 and with assessor 2 | Sensitivity vs assessor 1 | Specificity vs assessor 1 | Sensitivity vs assessor 2 | Specificity vs assessor 2 | PSA with assessor 1 | PSA with assessor 2 |
|---|---|---|---|---|---|---|---|
| Domain | GPT-6 Astra | 42.4 (0.0 to 64.1) [14/33] | 87.3 (74.2 to 97.4) | 43.3 (0.0 to 66.7) [13/30] | 87.0 (74.1 to 97.0) | 34.6 (0.0 to 58.1) | 33.3 (0.0 to 55.6) |
| Domain | Gemini 4 Argon | 33.3 (0.0 to 56.7) [11/33] | 95.1 (87.3 to 100.0) | 33.3 (0.0 to 59.6) [10/30] | 94.8 (86.5 to 100.0) | 38.6 (0.0 to 58.6) | 37.0 (0.0 to 62.9) |
| Overall | Second human assessor | 50.0 (21.7 to 73.5) [13/26] | 64.7 (37.5 to 83.0) | 52.0 (26.3 to 74.2) [13/25] | 62.9 (37.5 to 80.9) | 51.0 (23.8 to 73.7) | Same as previous column |
| Overall | RoB-BERT v6.1 | 57.7 (16.0 to 88.5) [15/26] | 73.5 (42.8 to 95.5) | 60.0 (15.8 to 89.7) [15/25] | 74.3 (41.7 to 97.7) | 60.0 (24.2 to 80.0) | 61.2 (18.2 to 82.6) |
| Overall | RoB-BERT reader alone | 57.7 (16.0 to 88.5) [15/26] | 73.5 (42.8 to 95.5) | 60.0 (15.8 to 89.7) [15/25] | 74.3 (41.7 to 97.7) | 60.0 (24.2 to 80.0) | 61.2 (18.2 to 82.6) |
| Overall | RoB-BERT encoder alone | 34.6 (0.0 to 70.6) [9/26] | 91.2 (69.0 to 100.0) | 36.0 (0.0 to 75.0) [9/25] | 91.4 (69.0 to 100.0) | 47.4 (0.0 to 75.0) | 48.6 (0.0 to 77.8) |
| Overall | Claude Fable 5.1 | 23.1 (0.0 to 57.1) [6/26] | 64.7 (30.0 to 100.0) | 24.0 (0.0 to 60.0) [6/25] | 65.7 (30.9 to 100.0) | 27.3 (0.0 to 50.9) | 27.9 (0.0 to 57.7) |
| Overall | GPT-6 Astra | 57.7 (16.0 to 88.5) [15/26] | 55.9 (22.2 to 90.0) | 60.0 (15.8 to 89.7) [15/25] | 57.1 (23.1 to 91.9) | 53.6 (19.0 to 75.3) | 54.5 (13.6 to 78.4) |
| Overall | Gemini 4 Argon | 46.2 (0.0 to 80.0) [12/26] | 82.4 (53.8 to 100.0) | 48.0 (0.0 to 81.8) [12/25] | 82.9 (52.9 to 100.0) | 54.5 (0.0 to 77.9) | 55.8 (0.0 to 81.8) |

Post hoc analysis not specified in the protocol. High is positive; low and some concerns are negative. An invalid output counts as not high. Values are % (95% CI); sensitivity cells also give [high-risk judgements matched / high-risk judgements by the reference assessor]. In the second-human rows, the first assessor's judgements are scored against the second assessor's and then the reverse. PSA, positive specific agreement, 2a / (2a + b + c), which does not depend on which assessor is treated as the reference. Human domain and overall judgements are each assessor's recorded judgements; system judgements come from the benchmark's RoB 2 algorithm, as in Table 4. Each system made one prediction per trial, reused across every assessor pair on that trial. Every pair includes at least one assessor who saw LLM suggestions. Counts are small (13 high-risk reference domains in panel a), so several intervals reach 0.0. Negative specific agreement and all cell counts are in the released analysis files.

### Supplementary Table S17. Sensitivity and specificity for a high-risk overall judgement, 419 pooled unassisted rows and subgroups.

**(a) Pooled unassisted rows (419 rows; 84 high-risk overall judgements in the reference).**

| AI system | Sensitivity for high-risk overall judgement, % (95% CI) [n/N] | Specificity, % (95% CI) | PSA, % (95% CI) | Overall judgements called high that the reference did not, n |
|---|---|---|---|---|
| RoB-BERT v6.1 | 52.4 (41.6 to 62.9) [44/84] | 70.1 (65.2 to 75.2) | 38.6 (30.2 to 46.6) | 100 |
| RoB-BERT reader alone | 53.6 (43.0 to 64.0) [45/84] | 69.9 (64.9 to 74.9) | 39.1 (30.7 to 47.1) | 101 |
| RoB-BERT encoder alone | 23.8 (15.1 to 32.5) [20/84] | 81.5 (77.2 to 85.5) | 24.1 (15.7 to 32.0) | 62 |
| Claude Fable 5.1 | 39.3 (29.3 to 50.5) [33/84] | 77.6 (73.3 to 82.1) | 34.4 (25.8 to 42.9) | 75 |
| GPT-6 Astra | 77.4 (67.9 to 85.7) [65/84] | 32.2 (27.3 to 37.5) | 34.6 (28.3 to 40.5) | 227 |

| AI system | Sensitivity for high-risk overall judgement, % (95% CI) [n/N] | Specificity, % (95% CI) | PSA, % (95% CI) | Overall judgements called high that the reference did not, n |
|---|---|---|---|---|
| Gemini 4 Argon | 39.3 (28.9 to 50.0) [33/84] | 84.5 (80.5 to 88.2) | 39.1 (29.4 to 48.0) | 52 |

**(b) By cohort, the ROBoto2 LLM-assisted rows, and the untruncated subset (SA3).**

| Population | AI system | Sensitivity for high-risk overall judgement, % (95% CI) [n/N] | Specificity, % (95% CI) | PSA, % (95% CI) | Overall judgements called high that the reference did not, n |
|---|---|---|---|---|---|
| ROBoto2 unassisted (n = 185) | RoB-BERT v6.1 | 65.7 (48.6 to 80.8) [23/35] | 65.3 (57.7 to 72.8) | 41.8 (29.5 to 52.2) | 52 |
| ROBoto2 unassisted (n = 185) | RoB-BERT reader alone | 68.6 (52.5 to 83.3) [24/35] | 65.3 (57.6 to 72.7) | 43.2 (30.6 to 53.5) | 52 |
| ROBoto2 unassisted (n = 185) | RoB-BERT encoder alone | 31.4 (16.7 to 48.0) [11/35] | 72.7 (65.1 to 80.0) | 25.3 (12.8 to 37.3) | 41 |
| ROBoto2 unassisted (n = 185) | Claude Fable 5.1 | 45.7 (29.2 to 62.2) [16/35] | 71.3 (63.8 to 78.6) | 34.0 (21.2 to 45.7) | 43 |
| ROBoto2 unassisted (n = 185) | GPT-6 Astra | 71.4 (55.2 to 85.7) [25/35] | 34.7 (27.2 to 42.3) | 31.6 (22.1 to 40.3) | 98 |
| ROBoto2 unassisted (n = 185) | Gemini 4 Argon | 37.1 (21.2 to 53.7) [13/35] | 79.3 (72.4 to 85.8) | 32.9 (19.5 to 46.0) | 31 |
| LogiMed (n = 192) | RoB-BERT v6.1 | 40.9 (26.8 to 55.3) [18/44] | 73.6 (66.2 to 80.9) | 35.6 (23.5 to 47.1) | 39 |
| LogiMed (n = 192) | RoB-BERT reader alone | 40.9 (26.8 to 55.3) [18/44] | 73.0 (65.3 to 80.1) | 35.3 (23.2 to 46.7) | 40 |
| LogiMed (n = 192) | RoB-BERT encoder alone | 20.5 (9.1 to 32.6) [9/44] | 86.5 (80.7 to 91.5) | 24.7 (11.4 to 37.1) | 20 |
| LogiMed (n = 192) | Claude Fable 5.1 | 36.4 (22.6 to 51.1) [16/44] | 80.4 (74.1 to 86.4) | 36.0 (23.3 to 48.5) | 29 |
| LogiMed (n = 192) | GPT-6 Astra | 81.8 (70.3 to 92.7) [36/44] | 31.1 (23.7 to 38.4) | 39.6 (30.5 to 48.5) | 102 |
| LogiMed (n = 192) | Gemini 4 Argon | 38.6 (24.1 to 53.5) [17/44] | 86.5 (80.7 to 91.7) | 42.0 (28.1 to 54.9) | 20 |
| Cochrane (n = 42) | RoB-BERT v6.1 | 60.0 (0.0 to 100.0) [3/5] ‡ | 75.7 (60.6 to 88.6) | 35.3 (0.0 to 63.2) | 9 |
| Cochrane (n = 42) | RoB-BERT reader alone | 60.0 (0.0 to 100.0) [3/5] ‡ | 75.7 (60.6 to 88.6) | 35.3 (0.0 to 63.2) | 9 |
| Cochrane (n = 42) | RoB-BERT encoder alone | 0.0 (0.0 to 0.0) [0/5] ‡ | 97.3 (91.2 to 100.0) | 0.0 (0.0 to 0.0) | 1 |
| Cochrane (n = 42) | Claude Fable 5.1 | 20.0 (0.0 to 66.7) [1/5] ‡ | 91.9 (81.8 to 100.0) | 22.2 (0.0 to 57.1) | 3 |
| Cochrane (n = 42) | GPT-6 Astra | 80.0 (33.3 to 100.0) [4/5] ‡ | 27.0 (13.2 to 41.7) | 22.2 (5.7 to 40.0) | 27 |
| Cochrane (n = 42) | Gemini 4 Argon | 60.0 (0.0 to 100.0) [3/5] ‡ | 97.3 (91.4 to 100.0) | 66.7 (0.0 to 100.0) | 1 |
| ROBoto2 assisted (n = 151) | RoB-BERT v6.1 | 50.9 (37.7 to 64.0) [27/53] | 68.4 (58.7 to 77.6) | 48.6 (36.7 to 59.3) | 31 |
| ROBoto2 assisted (n = 151) | RoB-BERT reader alone | 50.9 (37.7 to 64.0) [27/53] | 68.4 (58.7 to 77.6) | 48.6 (36.7 to 59.3) | 31 |
| ROBoto2 assisted (n = 151) | RoB-BERT encoder alone | 15.1 (5.7 to 25.4) [8/53] | 88.8 (82.4 to 94.8) | 22.2 (9.0 to 34.9) | 11 |
| ROBoto2 assisted (n = 151) | Claude Fable 5.1 | 32.1 (20.0 to 45.6) [17/53] | 78.6 (70.1 to 86.3) | 37.4 (24.7 to 50.0) | 21 |
| ROBoto2 assisted (n = 151) | GPT-6 Astra | 67.9 (55.2 to 80.4) [36/53] | 33.7 (24.2 to 43.2) | 46.8 (36.7 to 55.9) | 65 |
| ROBoto2 assisted (n = 151) | Gemini 4 Argon | 30.2 (17.9 to 42.9) [16/53] | 80.6 (72.0 to 88.2) | 36.4 (22.5 to 48.8) | 19 |
| Pooled unassisted, untruncated (SA3) (n = 332) | RoB-BERT v6.1 | 59.4 (48.0 to 71.1) [41/69] | 71.9 (66.4 to 77.2) | 44.6 (35.4 to 53.1) | 74 |

| **Population** | **AI system** | **Sensitivity for high-risk overall judgement, % (95% CI) [n/N]** | **Specificity, % (95% CI)** | **PSA, % (95% CI)** | **Overall judgements called high that the reference did not, n** |
|---|---|---|---|---|---|
| Pooled unassisted, untruncated (SA3) (n = 332) | RoB-BERT reader alone | 60.9 (49.4 to 72.6) [42/69] | 71.5 (65.9 to 76.9) | 45.2 (36.2 to 53.6) | 75 |

Post hoc analysis not specified in the protocol. Format follows Table 4. High is positive; low and some concerns are negative. An invalid output counts as not high. PSA, positive specific agreement. Intervals are percentile 95% intervals from 2,000 trial-level cluster-bootstrap resamples (seed 20261103), as in Table 4. ‡ The Cochrane cohort has only 5 high-risk reference overall judgements; in 10 of 2,000 resamples no high-risk reference judgement was drawn and sensitivity was undefined, so its intervals are very wide. SA3 is the pre-specified untruncated subset (332 of the 419 pooled unassisted rows), shown for RoB-BERT and the reader alone because truncation explained the LogiMed deficit.
